# Timing dependent synergies between motor cortex and posterior spinal stimulation in humans

**DOI:** 10.1101/2023.08.18.23294259

**Authors:** James R. McIntosh, Evan F. Joiner, Jacob L. Goldberg, Phoebe Greenwald, Lynda M. Murray, Earl Thuet, Oleg Modik, Evgeny Shelkov, Joseph M. Lombardi, Zeeshan M. Sardar, Ronald A. Lehman, Andrew K. Chan, K. Daniel Riew, Noam Y. Harel, Michael S. Virk, Christopher Mandigo, Jason B. Carmel

**Affiliations:** Dept. of Neurology; Dept. of Orthopedic Surgery; Dept. of Neurological Surgery, Columbia University, 650 W. 168th St, New York, NY, 10032, USA; Dept. of Neurological Surgery; Dept. of Neurology, Weill Cornell Medicine - New York Presbyterian, Och Spine, 1300 York Ave, New York, NY 10065; New York Presbyterian, The Och Spine Hospital, 5141 Broadway, New York, NY 10034; Departments of Neurology; Rehabilitation and Human Performance, Icahn School of Medicine at Mount Sinai, One Gustave L. Levy Place, New York, NY 10029; James J. Peters VA Med. Ctr., 130 West Kingsbridge Road, Bronx, NY 10468

**Keywords:** Epidural, Electrical stimulation, Cervical, Motor cortex, Spinal cord, Myelopathy, Motor evoked potentials

## Abstract

Volitional movement requires descending input from motor cortex and sensory feedback through the spinal cord. We previously developed a paired brain and spinal electrical stimulation approach in rats that relies on convergence of the descending motor and spinal sensory stimuli in the cervical cord. This approach strengthened sensorimotor circuits and improved volitional movement through associative plasticity. In humans it is not known whether dorsal epidural SCS targeted at the sensorimotor interface or anterior epidural SCS targeted within the motor system is effective at facilitating brain evoked responses. In 59 individuals undergoing elective cervical spine decompression surgery, the motor cortex was stimulated with scalp electrodes and the spinal cord with epidural electrodes while muscle responses were recorded in arm and leg muscles. Spinal electrodes were placed either posteriorly or anteriorly, and the interval between cortex and spinal cord stimulation was varied. Pairing stimulation between the motor cortex and spinal sensory (posterior) but not spinal motor (anterior) stimulation produced motor evoked potentials that were over five times larger than brain stimulation alone. This strong augmentation occurred only when descending motor and spinal afferent stimuli were timed to converge in the spinal cord. Paired stimulation also increased the selectivity of muscle responses relative to unpaired brain or spinal cord stimulation. Finally, paired stimulation effects were present regardless of the severity of myelopathy as measured by clinical signs or spinal cord imaging. The large effect size of this paired stimulation makes it a promising candidate for therapeutic neuromodulation.

## Key points summary

● Pairs of stimuli designed to alter nervous system function typically target the motor system alone or the sensorimotor convergence in cortex.
● In humans undergoing clinically indicated surgery we tested a paired brain and spinal cord stimulation paradigm that we developed in rats to target sensorimotor convergence in the cervical spinal cord.
● Arm and hand muscle responses to paired sensorimotor stimulation were six times larger than brain or spinal cord stimulation alone when applied to the posterior but not anterior spinal cord.
● Arm and hand muscle responses to paired stimulation were more selective for targeted muscles than the brain- or spinal-only conditions, especially at latencies that produced the strongest effects of paired stimulation.
● The paired stimulation effect was independent of the degree of myelopathy, suggesting that it could be applied as therapy in people affected by disorders of the central nervous system.

## 1 Introduction

Learning and execution of skilled movements, such as reaching and grasping, require coincident activity of motor and sensory circuits. Identifying how sensory and motor connections integrate has been an important goal for understanding movement (Wolpert, Diedrichsen, and Flanagan 2011). One tractable way to test and modify sensorimotor circuits has been to deliver pairs of exogenous stimuli (Asan, McIntosh, and Carmel 2022), one to the sensory and the other to the motor system. For example, paired associative stimulation (Stefan et al. 2000) combines motor cortex stimulation and sensory nerve stimulation at the wrist. Paired stimulation techniques provide the opportunity to probe nervous system interactions and to change connections through activity-dependent plasticity.

It is not known which sites of sensorimotor integration might be best to target with paired stimulation to strengthen circuits in health or disease. Paired associative stimulation targets the brain and can modulate muscle responses from motor cortex stimulation up or down depending on timing. Similarly, we hypothesised that timing motor cortex and spinal sensory stimulation to converge in the spinal cord would target another key site of sensorimotor integration. To test this, we have used epidural electrodes to pair motor cortex stimulation with dorsal (sensory) *spinal cord* stimulation in rats. Epidural spinal cord stimulation recruits a large number of sensory axons as they enter the spinal cord (Capogrosso et al. 2013; Liang et al. 2023; McIntosh et al. 2023). In one preclinical study, our group showed that timing motor cortex and dorsal epidural spinal stimulation to converge in the spinal cord markedly increased the motor evoked potential (MEP) (by 155%), while convergence in motor cortex resulted in a much smaller effect (17%) (Pal et al. 2022). We have shown this method of paired stimulation targeted at sensorimotor interactions in the spinal cord to induce plasticity (Mishra et al. 2017) and improve dexterity in rats with spinal cord injury (Pal et al. 2022).

Our sensorimotor approach contrasts with other successful paired stimulation approaches that target only the motor system. Foundational experiments paired stimuli within the motor cortex (Jackson, Mavoori, and Fetz 2006; Seeman et al. 2017) to induce plasticity. More analogous to our approach, paired stimulation has been used to target interactions between the corticospinal tract and motoneurons in the spinal cord. One approach used repetitive intraparenchymal motor cortex and spinal cord stimulation in non-human primates (Nishimura et al. 2013) to induce plasticity. In humans, corticospinal stimulation can be delivered non-invasively with transcranial magnetic stimulation while non-invasive peripheral nerve stimulation is used to activate motoneurons antidromically (Taylor and Martin 2009; Bunday and Perez 2012). These techniques have been demonstrated to induce plasticity in the motor system and to improve movement in people with spinal cord injury (Bunday, Urbin, and Perez 2018).

Other paired stimulation approaches have shown effects of convergent stimuli, but which circuits were activated was less clear. In humans, pairing brain stimulation with non-invasive spinal cord stimulation techniques (Roy, Bosgra, and Stein 2014; Knikou 2014; Al’joboori et al. 2021) and mixed motor and sensory peripheral nerve stimulation (Cowan et al. 1986; Deuschl et al. 1991; Poon et al. 2008; Guzmán-López et al. 2012), have been investigated. The ability to use more targeted epidural anterior or posterior spinal cord stimulation (Guiho, Baker, and Jackson 2021) in humans paves the way to test whether such targeting, either motor-motor or sensorimotor respectively, effectively facilitates motor responses to brain stimulation in humans.

Moreover, paired stimulation may enable greater selectivity of muscles that are commonly targeted by stimulation at either of the two sites. Electrical stimulation of the motor cortex can produce widespread activation across multiple muscles (Szelényi, Kothbauer, and Deletis 2007). Our previous work (McIntosh et al. 2023) showed that epidural spinal cord stimulation (SCS) produces the largest MEPs from muscles innervated by the stimulated spinal segment with spread to adjacent myotomes. Combined brain and spinal cord stimulation may allow an increase of selectivity for the muscles activated in common by both methods. Increased selectivity may allow equivalent strength of activation using lower stimulation intensity, thereby enhancing safety and tolerability.

We hypothesised that motor cortex and epidural cervical spinal cord stimulation would create strong and selective muscle responses when descending motor and spinal sensory stimuli converge in the cervical cord. We tested this hypothesis in the operating room in people undergoing clinically indicated elective spine surgery. We took advantage of the fact that spinal decompression surgery to relieve stenosis is done either via an anterior or a posterior approach. This enabled us to pair motor cortex stimulation with either spinal motor (anterior) or sensory (posterior) stimulation. This allowed us to extend our findings in rats to test the proper site of paired stimulation and whether pairing changes the strength or selectivity of arm and hand muscle activation.

## 2 Material and methods

### 2.1 Experimental design

We conducted a single session physiology study in people during clinically indicated surgery for cervical stenosis to address this hypothesis. Participants were identified prospectively by study personnel when they were scheduled for elective surgery with the goal of spinal cord decompression via either an anterior or posterior approach. Anterior surgery included removal of the intervertebral disc or the vertebral body, while posterior surgery included laminectomy. Both approaches provided access to the epidural space. To make stimulation as specific as possible we positioned the electrodes at the medio-lateral location where roots either enter (dorsal) or exit (anterior) the spinal cord. The primary outcome of the study was change in the MEP size recorded from arm muscles during paired stimulation compared to brain or spinal cord stimulation alone. To test the proper timing of stimulation, the inter-stimulus interval (ISI) between motor cortex and spinal stimulation was varied. To determine the differential effects of sensory versus motor spinal cord stimulation, suprathreshold motor cortex stimulation was combined with either dorsal (sensory) or anterior (motor) spinal cord stimulation performed below motor threshold (Supporting Information Table 1). To quantify whether pairing produces synergistic effects, suprathreshold brain stimulation and suprathreshold spinal stimulation were compared to each site alone. To further examine the role of timing, we paired single-pulse and subthreshold motor cortex stimulation with suprathreshold spinal stimulation. In order to determine the selectivity of paired stimulation for the targeted muscle, optimal pairing was compared to suprathreshold brain-only and suprathreshold spinal-only stimulation. Finally, to determine whether spinal cord and nerve root compression alter the effects of paired stimulation, the MEPs of paired stimulation were compared in segments with and without compression and also analysed in relation to clinical signs of neural injury.

**Table 1.** Summary clinical characteristics of all participants separated by posterior vs anterior spinal cord stimulation.

|  | All | Posterior* | Anterior* | p** |
| --- | --- | --- | --- | --- |
| Total: | 59, (100%) | 47, (100%) | 13, (100%) |  |
| Age*** |  |  |  | 0.489 |
| 30-49 | 6, (10.2%) | 4, (8.5%) | 2, (15.4%) |  |
| 50-64 | 22, (37.3%) | 19, (40.4%) | 4, (30.8%) |  |
| 65+ | 31, (52.5%) | 24, (51.1%) | 7, (53.8%) |  |
| Sex |  |  |  | 0.233 |
| F | 27, (45.8%) | 20, (42.6%) | 8, (61.5%) |  |
| M | 32, (54.2%) | 27, (57.4%) | 5, (38.5%) |  |
| mJOA |  |  |  | 0.394 |
| 1-12 | 15, (25.4%) | 11, (23.4%) | 5, (38.5%) |  |
| 13-15 | 28, (47.5%) | 22, (46.8%) | 6, (46.2%) |  |
| 16-18 | 16, (27.1%) | 14, (29.8%) | 2, (15.4%) |  |
| Axial neck pain |  |  |  | 0.971 |
| Y | 36, (61%) | 29, (61.7%) | 8, (61.5%) |  |
| N | 19, (32.2%) | 15, (31.9%) | 4, (30.8%) |  |
| U | 4, (6.8%) | 3, (6.4%) | 1, (7.7%) |  |
| Radiating pain |  |  |  | 0.899 |
| Y | 29, (49.2%) | 23, (48.9%) | 6, (46.2%) |  |
| N | 26, (44.1%) | 21, (44.7%) | 6, (46.2%) |  |
| U | 4, (6.8%) | 3, (6.4%) | 1, (7.7%) |  |
| Weakness |  |  |  | 0.808 |
| Y | 46, (78%) | 36, (76.6%) | 10, (76.9%) |  |
| N | 10, (16.9%) | 9, (19.1%) | 2, (15.4%) |  |
| U | 3, (5.1%) | 2, (4.3%) | 1, (7.7%) |  |
| Unsteady gait |  |  |  | 0.728 |
| Y | 38, (64.4%) | 30, (63.8%) | 9, (69.2%) |  |
| N | 21, (35.6%) | 17, (36.2%) | 4, (30.8%) |  |
| Hyperreflexia |  |  |  | 0.069 |
| Y | 30, (50.8%) | 26, (55.3%) | 4, (30.8%) |  |
| N | 22, (37.3%) | 15, (31.9%) | 8, (61.5%) |  |
| U | 7, (11.9%) | 6, (12.8%) | 1, (7.7%) |  |
| Sphincter Dysfunction |  |  |  | 0.312 |
| Y | 5, (8.5%) | 3, (6.4%) | 2, (15.4%) |  |
| N | 54, (91.5%) | 44, (93.6%) | 11, (84.6%) |  |
| Severe Foraminal Stenosis |  |  |  | 0.816 |
| Y | 41, (69.5%) | 32, (68.1%) | 9, (69.2%) |  |
| N | 15, (25.4%) | 12, (25.5%) | 4, (30.8%) |  |
| U | 3, (5.1%) | 3, (6.4%) | 0, (0.0%) |  |
| T2 Signal Change |  |  |  | 0.424 |
| Y | 45, (76.3%) | 36, (76.6%) | 9, (69.2%) |  |
| N | 12, (20.3%) | 9, (19.1%) | 4, (30.8%) |  |
| U | 2, (3.4%) | 2, (4.3%) | 0, (0.0%) |  |
| Racial category |  |  |  |  |
| Asian | 2, (3.4%) | 1, (2.1%) | 1, (7.7%) |  |
| Black or African American | 8, (13.6%) | 6, (12.8%) | 2, (15.4%) |  |
| More than one Race | 2, (3.4%) | 1, (2.1%) | 2, (15.4%) |  |
| Unspecified | 17, (28.8%) | 15, (31.9%) | 2, (15.4%) |  |
| White | 30, (50.8%) | 24, (51.1%) | 6, (46.2%) |  |
| Ethnic category |  |  |  |  |
| Hispanic | 5, (8.5%) | 5, (10.6%) | 1, (7.7%) |  |
| Not Hispanic | 43, (72.9%) | 33, (70.2%) | 10, (76.9%) |  |
| Unspecified | 11, (18.6%) | 9, (19.1%) | 2, (15.4%) |  |

\*A single participant took part in both anterior and posterior portions of the experiment. \*\*Wilcoxon Rank Sum test. No correction for multiple comparisons was applied. \*\*\*At the time of surgery. mJOA, modified Japanese Orthopaedic Association (mJOA) score. No - N, Yes - Y, Unknown/Unspecified - U. Severe foraminal stenosis is defined as no visible fat around the nerve root within the foramen. Severe foraminal stenosis and T2 signal change were assessed between C4 and T1 segments.

### 2.2 Participants

The study enrolled adult patients who required surgical treatment for cervical spondylotic myelopathy or foraminal stenosis. Patients were recruited from the clinical practices of spine surgeons at the Och Spine Hospital and Weill-Cornell NewYork-Presbyterian. The study protocol was conducted in compliance with the Declaration of Helsinki after it was reviewed and approved by the Institutional Review Boards of Columbia University Irving Medical Center and Weill Cornell Medicine (ClinicalTrials.gov: NCT05163639). We limited the extension of surgery for the purpose of experimentation to no longer than 15 minutes to minimise the risk of increased surgical and anaesthetic complications (Brendler 1968).

Enrollment criteria: Participants were recruited if their clinically indicated surgery provided access to the cervical epidural space. Participants were excluded if they had neck or chest stimulation devices (e.g., vagal nerve stimulation, cardiac pacemaker), epilepsy, a history of skull surgery with metal implants, cochlear implants, aneurysm clips or stents in neck or cerebral blood vessels, or evidence of skull shrapnel. Before surgery, written informed consent was obtained from all participants. Standard of care preoperative clinical assessments were conducted, including clinical MRI scans to assess the degree of foraminal stenosis and the extent and location of T2 signal hyperintensity within the spinal cord. Foraminal stenosis was defined as severe when no fat could be observed around the nerve root within the foramen. The severity of myelopathy in terms of motor and sensory dysfunction in the upper and lower extremities, as well as urinary dysfunction, was assessed using the Modified Japanese Orthopaedic Association (mJOA) scale (Benzel et al. 1991). Mild myelopathy is defined as mJOA scores from 15 to 17, moderate myelopathy with mJOA from 12 to 14, and severe myelopathy is mJOA scores from 0 to 11.

We powered the study based on the first 5 participants taking part in the main stimulation paradigm: suprathreshold motor cortex stimulation paired with subthreshold posterior spinal cord stimulation. To achieve >80% power on the comparison of whether facilitation was greater than 0%, 12 participants would be needed. Additional participants were subsequently recruited to investigate other conditions and enable correction for multiple comparisons in the ISI range 3-13 ms.

### 2.3 Electrical stimulation and recording

Following anaesthesia induction and during recording, only total intravenous anaesthesia without paralytics was used. The Cadwell IOMAX (Cadwell Inc., WA, USA) intraoperative monitoring system was used for recording and stimulation. Muscles were chosen for intramuscular electromyogram (EMG) per standard of care, with additional recordings from wrist muscles (Fig. 1A). Due to clinical monitoring and hardware constraints, not all muscles were recorded in each experiment. On average, 16.1 muscles were recorded per participant. Biceps, triceps, APB, ADM, TA and AH were consistently recorded ipsilateral to the site of spinal cord stimulation (> 98% of participants). Wrist muscles (ECR and FCR) were recorded in 69.5% of participants. The deltoid was typically recorded (93.2%) instead of trapezius (6.8%). MEP responses were recorded with needles placed in the muscles (Rhythmlink, SC, USA or Ambu, Denmark) at a sampling rate of 8. <u>3</u> kHz and band-pass filtered between 10 Hz and 2 kHz.

**Figure 1.**
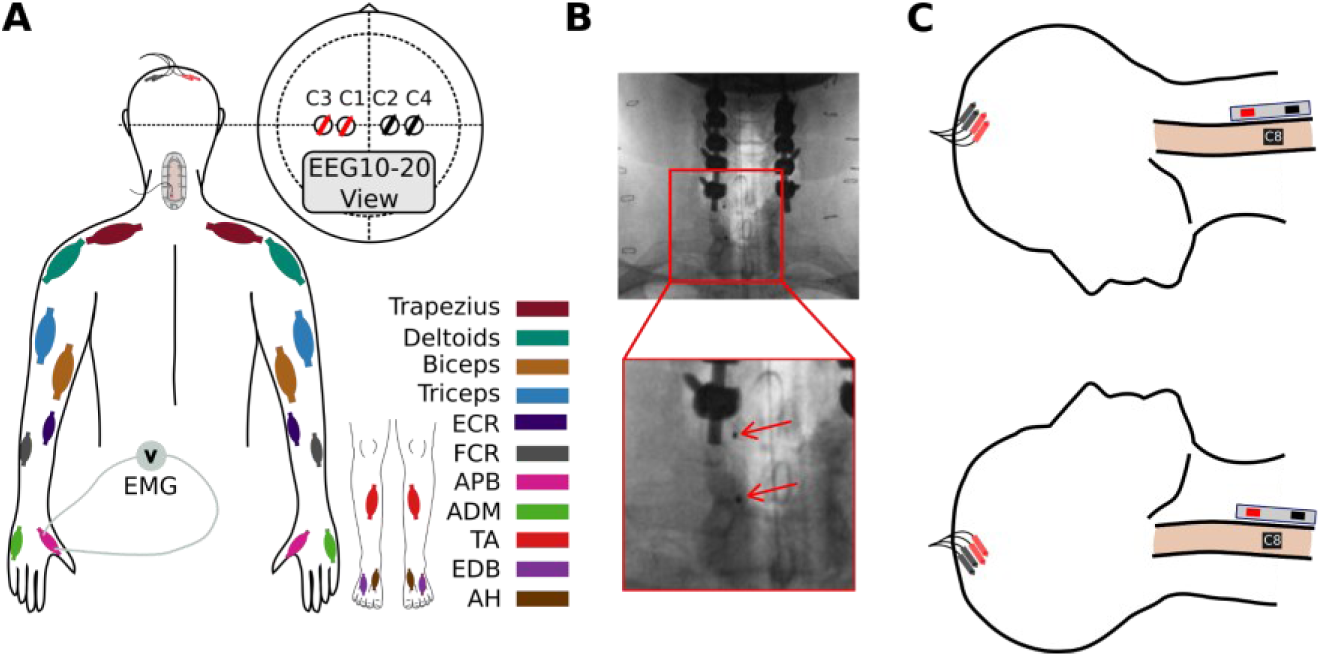
Epidural spinal cord and brain stimulation experiment during posterior and anterior cervical spine surgery. ***A***, Colors correspond to different recorded muscles (see legend). Subdermal needles were placed for brain stimulation. Catheter electrode shown placed below the lamina on the posterior aspect of the spinal cord. ***B***, Example of catheter placement targeting dorsal root fibres, relative to bony anatomy when the posterior aspect of the spinal cord is being stimulated. X-ray was acquired after surgical instrumentation but prior to removal of the catheter. Red arrows indicate the contacts of the catheter electrode. ***C***, The catheter electrode was used to stimulate the posterior (top) or anterior (bottom) aspect of the spinal cord in different participants. The posterior location targets the dorsal root entry zone, while the anterior location targets the ventral root exit.

Motor cortex stimulation was performed with transcranial electrical stimulation (tES). A monophasic triplet pulse with a width of 75μs and an ISI of 3ms was delivered through subdermal needle electrodes (Rhythmlink, SC, USA or AMC, FL, USA) placed in a quadripolar montage at C1, C2, C3, and C4 (see Fig. 1A inset). The two subdermal electrodes in the hemisphere targeted for stimulation served as the anode, and the two electrodes on the opposite hemisphere served as the cathode. The quadripolar configuration resulted in lower thresholds than bipolar stimulation in pilot experiments (data not shown). Triple-pulse stimulation can reduce MEP threshold, which is important for experiments during ongoing surgery, but the timing of descending activation is ambiguous. Hence, we employed single-pulse tES in a subset of experiments to determine precise timing of brain-spinal cord interactions. For experiments in 11 participants we observed reliable MEPs at 50V, the lowest stimulation intensity allowed by the intraoperative hardware. In these instances, in order to measure the MEP threshold we reduced the pulse width to 50μs for the duration of the experiment.

The experimental procedure began once the dura was exposed and the epidural stimulation electrode was positioned. For spinal cord stimulation (SCS), a single biphasic pulse (pulse-width = 250 μs) using a flexible catheter electrode with 1.3-mm contacts and 15-mm spacing (Ad-Tech Medical Instrument Corp, WI, USA) was used. Epidural electrodes were oriented in the rostro-caudal direction (Fig. 1B-C), with the cathode caudal and straddling the dorsal root entry zone (DREZ) or ventral root exit. The electrodes were positioned on the less affected side as determined by the muscle strength grading. If the strength grading was the same on both sides, the side with a lower threshold cortical-evoked MEPs was targeted. If the selected side was surgically inaccessible, the experiment was carried out on the accessible side. The location of the exiting or entering cervical nerve was estimated based on bony and neural anatomical landmarks. This method of hand placement of the electrode was previously validated by co-registering intraoperative computer tomography with pre-operative MRI and verifying the electrode placement relative to the DREZ (McIntosh et al. 2023). In a subset of cases (n = 23), intraoperative imaging was required as part of the surgical procedure and this was subsequently utilised to confirm the location of the electrode contacts (e.g., Fig. 1C).

#### 2.3.1 Determination of MEP Threshold and Choice of Target Muscle

Upon insertion of the cervical epidural spinal cord electrode, we determined the threshold for spinal-evoked MEPs by increasing the stimulation intensity in 0.5 mA steps. Next, we conducted an equivalent procedure for tES by incrementing the voltage in steps of 5 V and observing the threshold for cortical MEPs in multiple ipsilateral muscles. All trains of brain and spinal stimulation stimuli were delivered at a rate of 0.5 Hz. For both spinal- and brain-evoked MEPs, we defined threshold as the lowest intensity at which a response was present in at least 50% of trials. Spinal stimulation was confirmed to be subthreshold by recording five MEPs.

Candidate target muscles were those where both brain- and spinal-evoked MEPs could be observed at stimulation intensities that did not interfere with ongoing surgery, irrespective of which cervical segment was stimulated. We prioritised hand muscles, starting with the abductor pollicis brevis (APB), followed by the wrist, upper arm, and shoulder. Ultimately, we selected a single muscle as the target and re-determined thresholds to a resolution of 0.1 mA for spinal stimulation intensity and 1 V for tES intensity.

#### 2.3.2 Comparison of Facilitation of Cortical MEPs by Posterior vs Anterior Spinal Stimulation

To determine whether posterior or anterior spinal stimulation augments cortical MEPs, equivalent experiments were performed with epidural electrodes placed either in the posterior or anterior epidural space. Posterior spinal stimulation was targeted to the DREZ, and the ventral stimulation was targeted to the root exit zone of the segment exposed during the surgery. In a single participant, both anterior and posterior positioning was performed within the same surgery. Paired stimulation performed with triple-pulse tES was delivered at 110% of the MEP threshold, and subthreshold spinal stimulation at 90% of the MEP threshold. The ISI between the initiation of tES and subsequent SCS was varied between 3 and 13 ms. Each pairing event was repeated five times, with the MEPs sequentially averaged and saved for subsequent analysis. In a subset of experiments, pairing events were repeated ten times.

To quantify the size of the paired brain and spinal stimulation effect, the MEP size at each ISI was divided by the brain-only stimulation MEP. We defined the optimal ISI as the ISI at which the largest MEP was generated.

#### 2.3.3 Estimation of Synergy in Corticospinal Convergence

When applying tES and SCS each at 110% of MEP threshold, a simple additive model would predict that the resultant MEP is the sum of the brain-only and spinal-only MEPs. On the other hand, a synergistic model of the interaction of brain and spine stimulation would predict a resultant MEP that is greater than the sum of its parts. In order to determine whether the convergence of brain and spinal stimulation was additive or synergistic, we performed triple-pulse suprathreshold tES (110%) combined with suprathreshold SCS (110%), targeting either the DREZ or the ventral root exit zone.

Lastly, to investigate the effect of tES intensity on facilitation, we applied suprathreshold SCS in conjunction with subthreshold tES, where the intensity of tES was varied between 50V and its previously determined maximum.

#### 2.3.4 Estimation of the Precise Inter-stimulus Interval

To accurately determine the ISI required for SCS to produce convergence relative to tES, we employed single-pulse tES. In most cases, the cortical MEP threshold was above the maximum tested voltage (typically 300 V) or at intensities that would disrupt the ongoing surgical procedure. As a result, we used the maximum possible voltage that would neither interfere with surgery nor exceed 300 V. In cases where the threshold was observable, we maintained consistency with the subthreshold tES approach by setting the stimulation intensity to 90% of threshold. For the pairing condition, we combined subthreshold tES with suprathreshold SCS set at 110% to establish a baseline. Pairing was conducted with ISIs ranging from 0 ms to 5 ms.

#### 2.3.5 Epidural spinal cord recording

In order to directly determine the timing of corticospinal transmission in a single participant, tES was ramped from 50V to 300V while the catheter electrode was switched to ports for recording of electrophysiological potentials. The recorded potentials were bandpass filtered between 0.3 kHz and 10 kHz.

### 2.4 Data analysis

The intraoperative monitoring software’s data was exported to MATLAB (R2022a, MathWorks, Inc., MA, USA). MEPs were zero-phase filtered using a Butterworth design, with a fifth-order lowpass filter at a 500 Hz passband and a sixth-order bandstop filter with a 59–61 Hz stopband. The AUC was calculated over a window from 8.5 ms to 75 ms after the first stimulation pulse began in order to capture the range of brain and spinal MEP across multiple muscles. Study data were collected and managed using REDCap (Research Electronic Data Capture) electronic data capture tools hosted at Weill Cornell Medical Center and Columbia University Irving Medical Center (Harris et al. 2009; 2019).

#### 2.4.1 Statistical analysis

Values are reported as mean ± standard error of the mean (SE) except when the median is employed. Nonparametric statistical tests are used throughout (Wilcoxon rank-sum, signed-rank tests and Kruskal–Wallis tests, α = .05), and Bonferroni correction was applied unless otherwise noted.

#### 2.4.2 Artefact rejection

We employed the same method for rejecting MEPs as previously described (McIntosh et al. 2023). Briefly, rejection was based on principal component analysis and human observer confirmation. The process involved computing principal components for a specific muscle across multiple stimulation intensities, regressing them with each MEP, and ranking the responses based on the root mean square of the regression error. While blinded to the stimulation condition, a manually adjusted sliding scale was then applied to reject traces that did not appear physiological under visual inspection: deflections in baseline, spread of stimulation artefact into the evoked response, excessive line noise, and fluctuations that were not time-locked to other responses. These were typically related to electrocautery or drilling and appeared stereotypical; they were randomly distributed across different phases of the experiment. This led to 52,048 analysed MEPs of which 11,337 (21.8%) were rejected. When considering the targeted muscles of the arm and hand, 30,188 MEPs were analysed of which 2,949 (9.8%) were rejected. Note that this is higher than in our previous work with SCS (McIntosh et al. 2023) due to the tES induced artefact being more extensive and the presence of additional sources of artefact induced by the ongoing surgery. Rejection was performed blinded to the ISI.

#### 2.4.3 Calculation of pairing effect

To assess facilitation, we normalised the AUC at each tested pairing ISI using the following approach:

1. Suprathreshold tES paired with subthreshold SCS was divided by the suprathreshold brain-only stimulation after confirming the absence of spinal MEPs.
2. Subthreshold tES paired with suprathreshold SCS was divided by suprathreshold spinal-only stimulation after confirming the absence of tES MEPs.
3. Suprathreshold tES paired with suprathreshold SCS was divided by the sum of the suprathreshold spinal-only and suprathreshold brain-only stimulation (minus background activity (Guiho et al., 2021)).

#### 2.4.4 Across participant averaging

Normalised AUCs were converted to percentage facilitation and illustrated as bar charts. A facilitation of 0% indicates that pairing produces no change relative to baseline, and 100% indicates a doubling of the MEP size. To generate the across-participant average, AUCs were averaged among all participants that had experiments with a given condition. In cases where ISIs within our test range (3-13 ms) were missing due to experimental error, surgical procedure constraints, or presence of artefact, we applied linear interpolation (14% of data) at the individual participant level before calculating the average. Analyses without interpolating data did not change the major findings.

#### 2.4.5 Calculation of selectivity

To examine whether the combination of brain and spinal cord stimulation can isolate individual muscles more effectively than either brain-only or spinal-only stimulation, we conducted a comparison of selectivity across these conditions. The pairing MEPs used for this analysis were extracted from suprathreshold tES (110%) and subthreshold SCS (90%) conditions at the optimal ISI. For the brain-only condition, suprathreshold stimulation (110%) was used. We included only those cases in which suprathreshold (110%) spinal-only stimulation intensity was also tested, as this was used to estimate spinal-only selectivity values. This analysis uses all muscles recorded ipsilateral to spinal cord stimulation; however, because the intensity can only be set for a single targeted muscle, non-targeted muscles may be above or below threshold.

The selectivity for a particular muscle was calculated as its AUC divided by the sum of the AUCs of all other muscles recorded on the same side of the body (McIntosh et al. 2023). This calculation ensures that the sum of selectivities across all muscles equals one. The AUC of the target muscle was selected to compare the levels of selectivity in a given experiment. However, we also established a measure of selectivity that could be evaluated across muscles regardless of the target. For this across-muscle measure, we utilised the equation 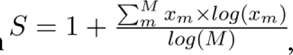, where is the individual muscle selectivity for muscle , and is the number of muscles. This equation incorporates the same structure as the entropy equation which has been previously used as a measure of selectivity (Lehky, Sejnowski, and Desimone 2005) but is normalised so that it takes a value of 0 when individual muscle selectivities are equal in all muscles and a value of 1 when only one muscle is activated.

There is likely to be an interaction between the size of the pairing effect and the selectivity of muscle recruitment. To determine this interaction, selectivity for all muscles was computed for each ISI. To determine the effect of pairing, the selectivity during brain and spinal cord pairing was divided by the selectivity of the summed suprathreshold tES (110%) and subthreshold SCS (90%) conditions.

#### 2.4.6 The impact of impairment on synergistic effects

To determine if paired stimulation was affected by nervous system injury, the degree of facilitation was analysed in relation to clinical and radiographic evidence of compression. In this analysis, the maximum pairing effect was compared with 3 pre-operative clinical scores related to myelopathy (strength, reflex, and mJOA) and radiographic evidence of neural injury (T2 signal hyperintensity and foraminal stenosis). Medical Research Council strength scores (Compston 2010) were averaged over the muscles of the forearm and hand on the stimulated side as these were the primarily targeted muscles. Reflex scores were derived from biceps and triceps on the targeted side as muscles of the forearm and hand were not typically tested. The maximum facilitation of suprathreshold brain and subthreshold spinal stimulation was used to summarise the strength of facilitation. Because of the strongly non-gaussian distribution of facilitation across participants, values were log transformed. Non-parametric Kruskal–Wallis one-way analysis of variance was used to assess the presence of a relationship between each of the dependent variables and the pairing facilitation strength. No correction for multiple comparisons was applied.

## 3 Results

### 3.1 Participant recruitment and characteristics

The study enrolled adult patients who required surgical treatment for cervical spondylotic myelopathy or multilevel foraminal stenosis (n = 63, 34M/29F, mean age 66 years, standard deviation = 11). In a subset of participants (n = 4), experimental procedures could not be attempted due to surgical constraints, and these participants have been excluded from further analysis. tES and SCS, at either the posterior (n = 46, 27M/19F), anterior (n = 12, 5M/7F) or both (n = 1, 1F) aspects of the cervical enlargement were performed. We determined that participants undergoing stimulation of the posterior aspect of the spinal cord were not detectably different from those undergoing stimulation of the anterior aspect in their demographics or degree of impairment (Table 1 and Supporting Information Table 2).

### 3.2 Posterior SCS augments motor cortex MEPs at the predicted convergence time, while anterior SCS does not

We observed strong facilitation between appropriately timed transcranial electrical stimulation (tES) and posterior SCS targeting the dorsal root entry zones (DREZ). By calculating the difference between brain-only and spinal-only MEP onset times, we predicted that convergence would occur at 9.8±0.6ms (Fig. 4). When suprathreshold tES was paired with subthreshold SCS (Fig. 1A-B), appropriate ISI for convergence in the spinal cord resulted in a significant facilitation of cortical MEPs in individual participants (Fig. 2). The degree of facilitation, when averaged across participants, was substantial, with an optimum ISI of 9 ms yielding an average increase of 577 ± 173% (*p* = 3.2×10^-5^, n = 38, signed-rank test; Fig. 3A). Facilitation was strongly time-dependent (*p* = 0.001, H(10) = 28.7, Kruskal–Wallis test), with the optimal ISI not different from the predicted convergence time (Fig. 4).

**Figure 2.**
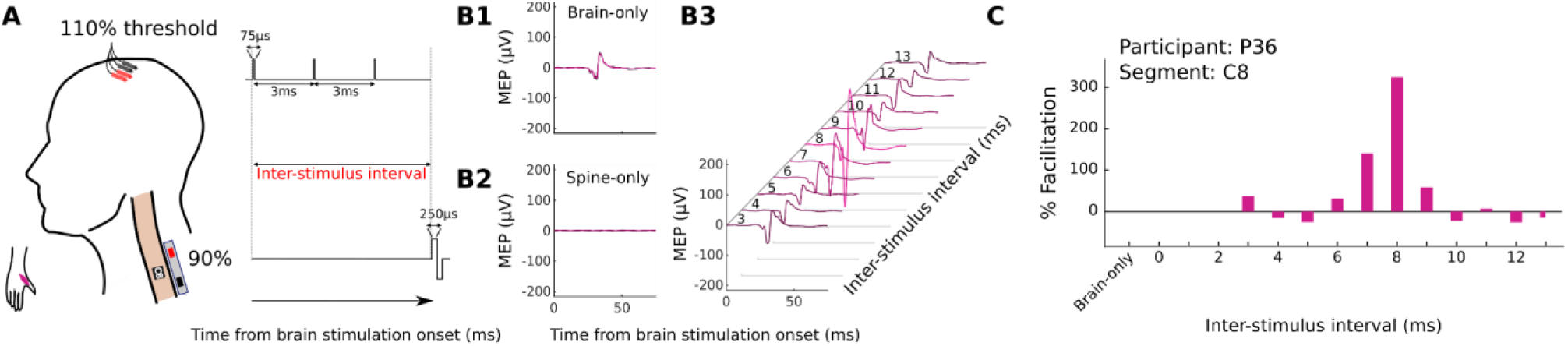
Experimental paradigm and results of varying the timing of spinal stimulation relative to transcranial electrical stimulation (tES) in a single participant. *A*, Schematic: three pulses are delivered over the motor cortex followed by a variable period of time (inter-stimulus interval; ISI) before a single pulse is delivered to the spinal cord. The catheter electrode was positioned over the C8 dorsal spinal cord, and the abductor pollicis brevis (APB) was the target muscle. *B1*, Brain-only baseline condition. The intensity of cortical stimulation was set to 110% of the APB threshold, ensuring a small MEP in the brain-only condition. **B2,** Spinal baseline condition. The intensity of spinal stimulation was set to 90% of the APB threshold, so no MEP was observed with spinal-only stimulation. *B3,* Paired stimulation. Averaged responses over 5 trials with variable ISI. *C*, Quantification of pairing facilitation. The facilitation is calculated relative to the brain-only MEP size. Facilitation of 324% was observed when the inter-stimulus interval was set to 8 milliseconds.

**Figure 3.**
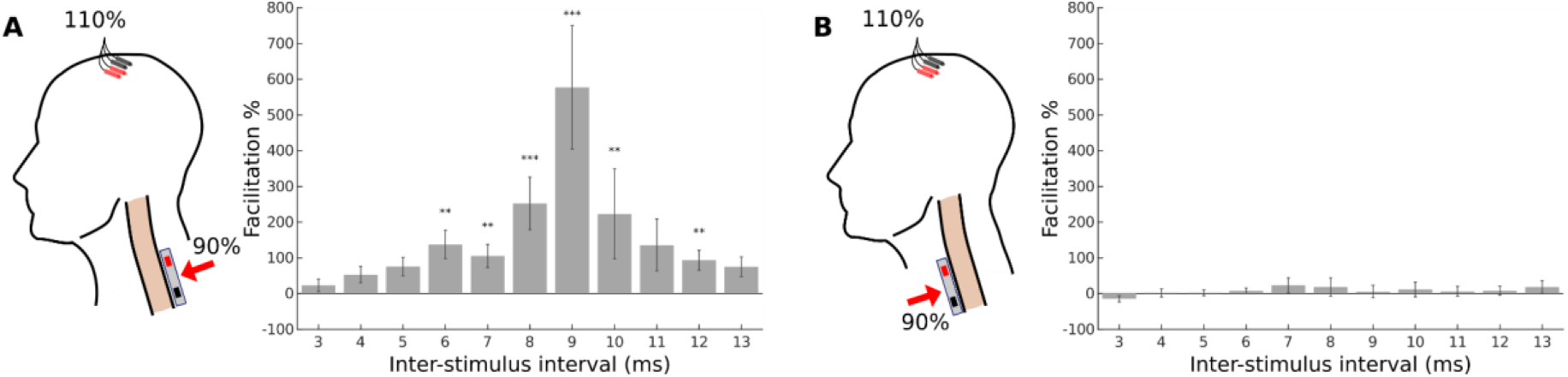
Augmentation of motor cortex MEPs with posterior, but not anterior, spinal stimulation. *A*, Schematic: 110% threshold transcortical electrical stimulation is combined with 90% threshold posterior cervical spinal stimulation. A strong facilitation is present when averaging across participants (n = 38, 23M/15F). ***B***, Schematic: as in A but cervical stimulation applied to the anterior aspect of the spinal cord. Anterior stimulation results in no observable facilitation (n = 12, 4M/8F). Across-participant signed-rank test, \**p* < .05, \*\**p* < .01, \*\*\**p* < .001, Bonferroni corrected for multiple comparisons.

**Figure 4.**
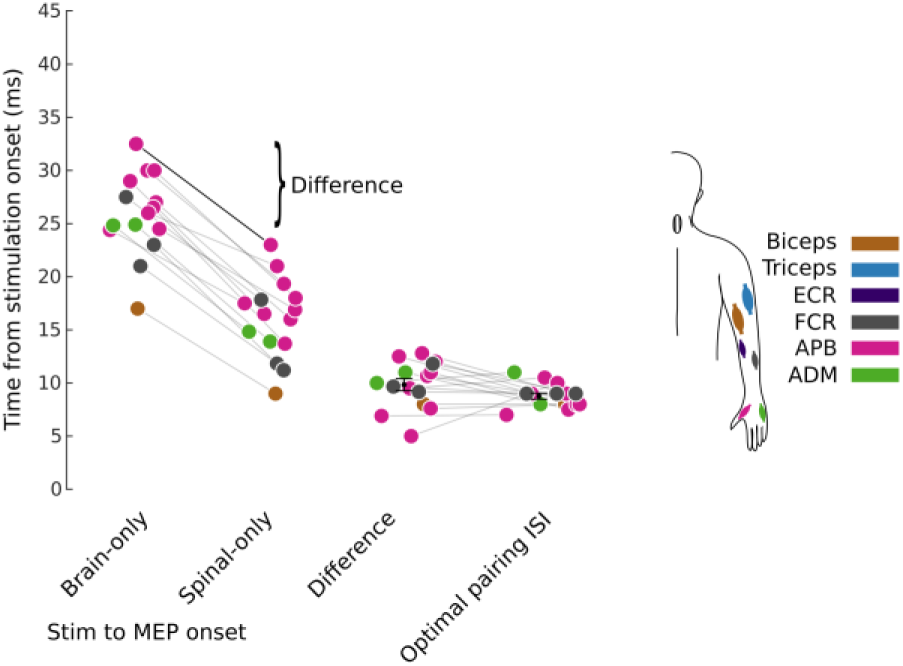
Estimate of optimal pairing ISI from brain and spinal MEP onset times. Subtracting the brain-only MEP onset time from the spinal-only MEP onset time produces a difference in the onset times (9.8±0.6ms) which acts as an estimate of the spinal cord convergence time. This estimate is not significantly different from the estimate of the optimal pairing ISI (8.8±0.3ms; p = 0.15, signed-rank test). Onset times for brain-only and spinal-only MEPs were estimated programmatically and refined manually. The programmatic detection extracted the first time point where the MEP magnitude was nine times its pre-stimulation standard deviation. This time point was then further refined by tracing the MEP back to its x-axis crossing. The optimal pairing ISI was estimated by taking the time of maximum facilitation for all cases where facilitation was greater than 50%. Connecting lines represent the same participant. Marker colours correspond to targeted muscles. Data shown only for participants (n = 15, 9M/6F) where an estimate of the brain-only, spinal-only and optimal pairing ISI could be made.

In contrast, when anterior spinal cord stimulation was paired with tES, there was no facilitation (23 ± 21% at an optimum ISI of 7 ms, *p* = 1.0, n = 12, signed-rank test; Fig. 3B). A direct comparison of the maximum facilitation of each participant between posterior (Supporting Information Fig. 1) and anterior (Supporting Information Fig. 2) stimulation demonstrated that posterior stimulation was more effective than anterior stimulation (*p* = 0.006, nposterior = 38, nanterior = 12, Wilcoxon Rank Sum test).

### 3.3 Synergistic effects of brain and spinal stimulation

To better understand the interactions of brain and spinal stimulation, we stimulated each site above motor threshold. We hypothesised there would be synergistic effects (i.e. the combined effects would be much larger than the MEPs of brain- or spinal-only stimulation). Using suprathreshold (110%) tES and posterior SCS, we altered the relative timing of stimulation.

A direct comparison of posterior and anterior stimulation demonstrated that suprathreshold posterior SCS was more effective than suprathreshold anterior SCS (*p* = 0.034, nposterior = 10, nanterior = 8, Wilcoxon Rank Sum test). The optimal ISI was 9 ms (n = 10; Fig. 5A), similar to Fig. 3. The facilitation at this ISI was strongly synergistic (average = 1166 ± 537% relative to the sum of brain-only and spinal-only MEPs) but did not reach statistical significance (*p* = 0.16, n = 10, signed-rank test, lowest *p* = 0.02 at 8ms not significant after correction for multiple comparisons). In contrast, with anterior SCS there was no facilitation, and the MEPs were simply additive (average = 31 ± 29% at peak ISI of 11 ms, *p* = 1.0, n = 8, signed-rank test; Fig. 5B).

**Figure 5.**
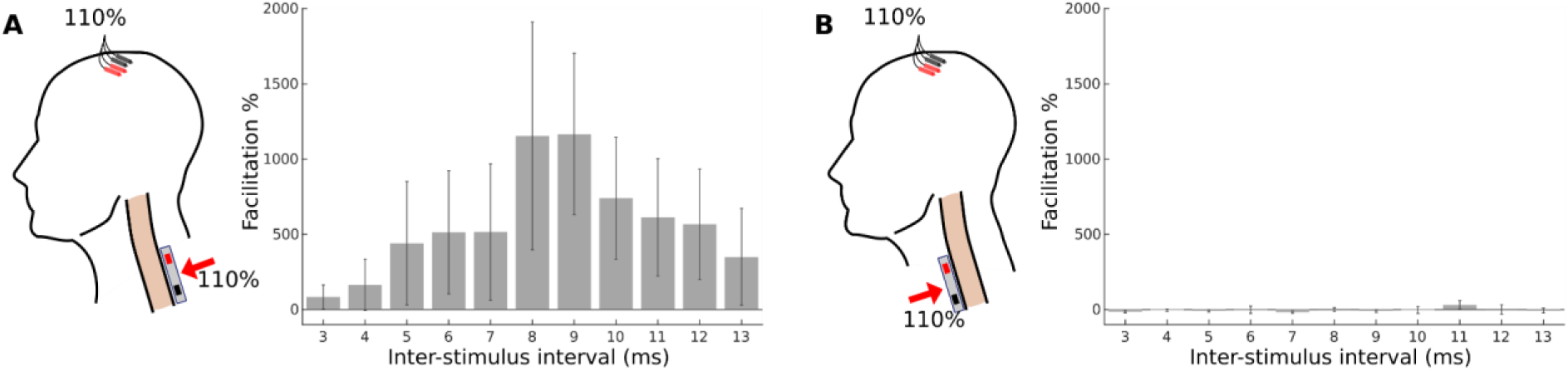
Suprathreshold posterior but not anterior spinal cord stimulation produces synergistic effects when paired with suprathreshold tES. *A*, Schematic: 110% threshold transcortical electrical stimulation is combined with 110% threshold posterior cervical spinal stimulation. A strong facilitation is present when averaging across participants (n = 10, 6M/4F). The peak facilitation is 1174% at 9ms relative to the sum of brain-only and spinal-only stimulation. *B*, Schematic: as in A but cervical stimulation applied to the anterior aspect of the spinal cord. In contrast, posterior aspect stimulation anterior stimulation results in no observable facilitation (n = 8, 3M/5F; peak facilitation = 38% at 11 ms).

### 3.4 The timing of brain and spinal stimulation convergence

To better understand the timed interactions between brain and spinal stimulation, we varied the time between a single pulse of tES and SCS (Fig. 6A). In the previous experiments, the interpretation of the timing of facilitation, as displayed in Fig. 3 and Fig. 5, was complicated by the application of three consecutive tES pulses over a span of 6ms, which was needed to reliably evoke a cortical MEP. Single pulse tES, as depicted in Fig. 6, does not typically generate an MEP at the voltages we employed (less than 300 V). However, given our previous observations of pronounced amplification in spinal circuits, we hypothesised that facilitation would still occur if suprathreshold SCS were introduced with an appropriate ISI relative to the subthreshold single pulse tES.

**Figure 6.**
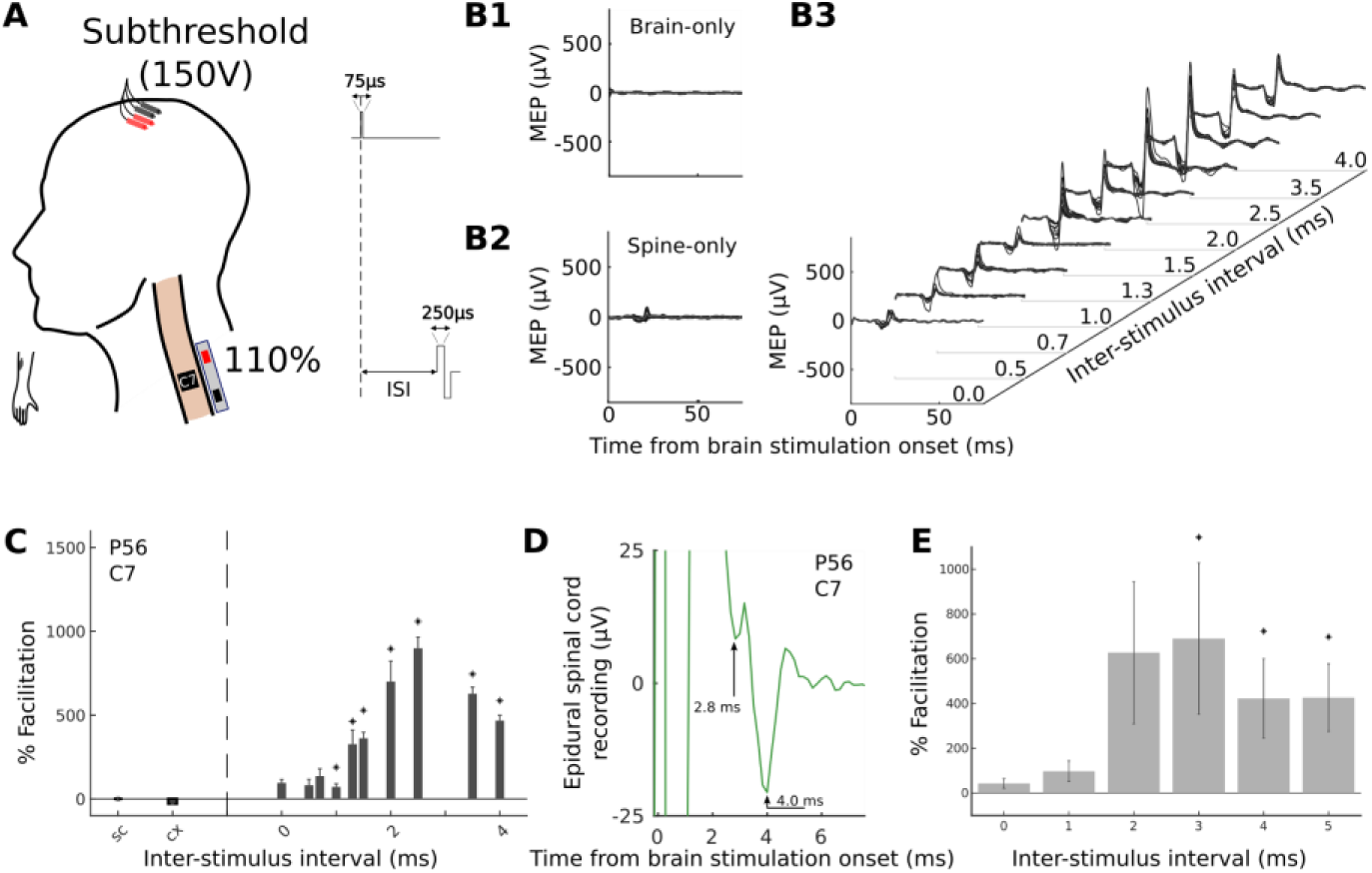
Facilitation occurs 2-3ms after cortical stimulation. *A*, Schematic: Subthreshold (150V) single pulse transcortical electrical stimulation is combined with 110% threshold posterior cervical spinal stimulation. Example for an individual participant (P56). The catheter electrode was positioned over the C7 dorsal root entry zone (DREZ) of the spinal cord and the FCR was the target muscle. ***B1***, Brain-only baseline condition. The intensity of transcranial stimulation was set to 150 V and no MEP was present below the maximum tested 300 V. ***B2,*** Spinal-only baseline condition. The intensity of spinal stimulation was set to 110% of the target threshold needed to induce a motor evoked potential (MEP). ***B3***, Paired stimulation. Averaged responses over 10 trials with variable ISI. ***C***, Quantification of pairing facilitation. The facilitation is calculated relative to the spinal-only MEP size. While the peak facilitation appears to be at 2.5 ms, the earliest facilitation appears to be in the range 1-1.5 ms. ***D***, Epidural spinal recordings. Brain-only stimulation was applied while a recording was made from the spinal electrode. A deflection is visible starting at 2.8 ms with the maximal deflection occurring at 4 ms. The stimulation artefact prior to 2.5 ms has been clipped for visualisation purposes. ***E***, Average over participants (n = 11, 4M/7F) receiving subthreshold (77-288 V) single pulse transcortical electrical stimulation combined with 110% threshold posterior cervical spinal stimulation. The optimal ISI when single pulse cortical stimulation is used is 3ms.

In an individual participant we evaluated ISIs at sub-millisecond precision to determine the onset and optimal ISI of facilitation (Fig. 6C-E). We also measured the recruitment curve of tES intensity at the optimal ISI to determine the dynamics of facilitation (Fig. 7). Additionally, epidural spinal cord recordings in response to tES were made to assess the transmission time from the brain to the spinal cord (Fig. 6E).

**Figure 7.**
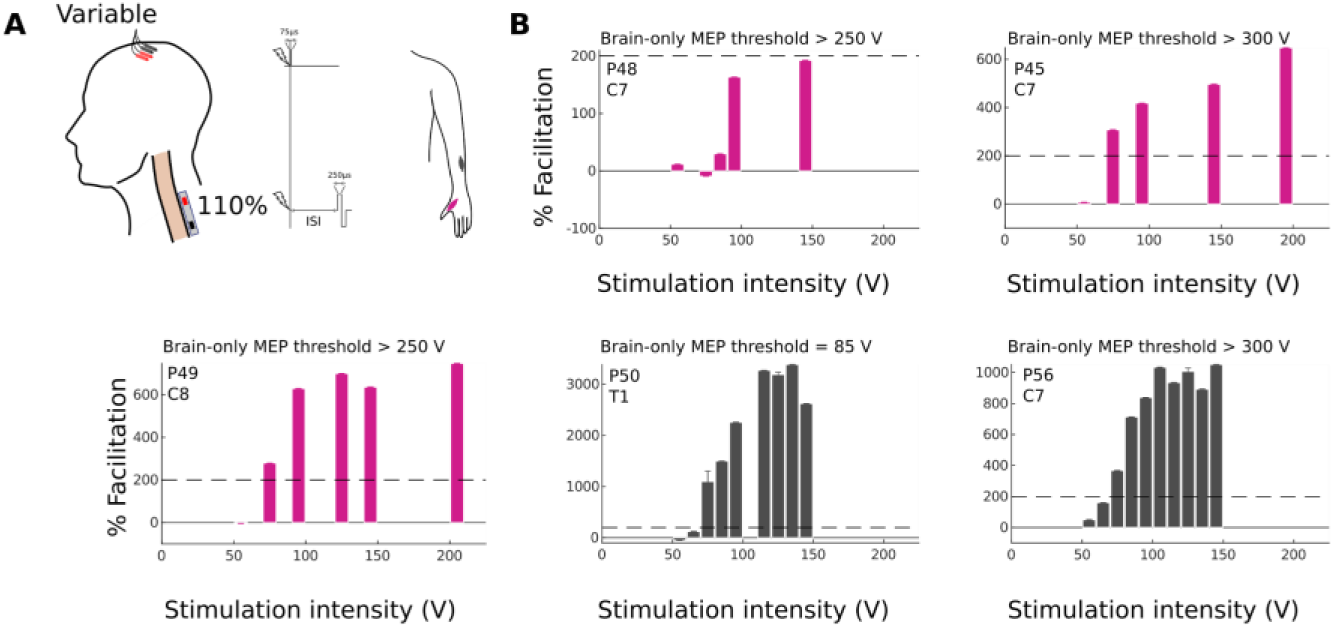
Suprathreshold spinal MEPs are strongly facilitated by extremely subthreshold single pulse transcranial electrical stimulation. *A*, Schematic: a single pulse delivered to the brain and spinal cord and choice of stimulation intensities. *B*, Cortical stimulation intensity was adjusted upwards from 50V while spinal stimulation intensity was maintained at 110% of threshold (n = 5, 1M/4F). Pairing was applied at the optimal inter-stimulus interval as determined in a previous experiment. Facilitation is initiated between 50V and 100 V in all cases, which is considerably lower than the threshold for brain-only stimulation in the majority of experiments (see text in figures). Bar colours correspond to the targeted muscle as shown in A.

Facilitation began to be observed with spinal stimulation initiated between 1.0 ms and 1.3 ms after brain stimulation (Fig. 6A-C) and reached its maximum at an ISI of 2.5 ms (899%, *p* = 0.018, n = 10 repetitions, signed-rank test). Notably, even without a detectable cortical MEP in the target muscle at voltages up to 300 V for this participant, facilitation initiated within the 50-75 V range (Fig. 7B, P56) and appeared to saturate at roughly 100 V when an ISI of 2ms was used.

Epidural spinal cord recordings of tES also referred to as D-waves (Vedran Deletis, Sala, and Ulkatan 2012) were made in the same participant as an indication of the corticospinal transmission time. The initial deflection is visible starting at 2.8 ms after the initiation of stimulation at the intensity used for pairing brain and spine.

We confirmed that the facilitation was present in the 2-3 ms range by pairing single pulse tES and SCS in a further 10 participants (Fig. 6E). Maximum facilitation was found to be 690.5 ± 338% at 3.0 ms (*p* = 0.041, n = 11, signed-rank test; Bonferroni corrected in the tested interval 0-5 ms). This optimal interstimulus interval is consistent with paired triple-pulse tES stimulation generating the largest MEP 3 ms after the last tES pulse (Fig. 3, Fig. 5). Further, this demonstrates that subthreshold brain stimulation can augment suprathreshold spinal stimulation.

### 3.5 Synergy with tES far below motor threshold

Facilitation of spinal MEPs was present even when combined with tES far below motor threshold. In five experiments we investigated the dependence of the paired facilitation to the stimulation intensity (Fig. 7A). At the optimal ISI using high intensity single-pulse tES (Fig. 7B), we stimulated the spinal cord at 110% of the MEP threshold while ramping up the single pulse tES intensity from 50V. Facilitation was initiated between 50 V and 100 V in all cases, which was considerably lower than the threshold for brain-only single-pulse stimulation in the majority of experiments (>250 V).

### 3.6 Paired stimulation activated muscles more selectively

Synergistic effects of stimulation were greatest in muscles targeted by brain and spinal stimulation in two representative cases (Fig. 8). In each, the tES was performed at 110% of threshold and SCS at 90% of threshold. In Fig. 8B, the spinal cord electrode was placed at C6, and tES intensities were optimised for the biceps muscle. While the strongest facilitation is presented in the biceps (1850%), it is also apparent in triceps (190%). In Fig. 8C, the spinal electrode was positioned at C8, and tES was optimised for the APB muscle. The strongest facilitation is present in the APB (138% increase).

**Figure 8.**
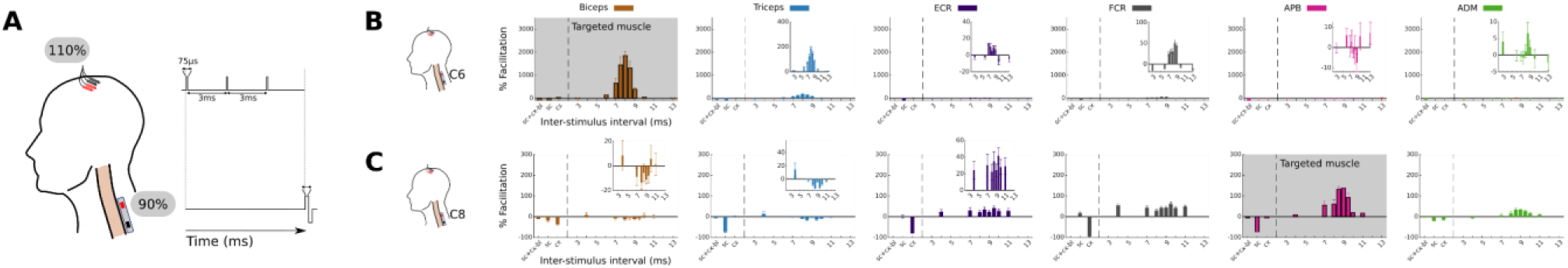
Facilitation is greatest in targeted muscles. *A*, Schematic: a triple pulse stimulation delivered to the brain and single pulse stimulation delivered to the spinal cord. For the muscle that was optimised for, intensity of brain stimulation was set to be 110% of threshold, and the intensity of spine stimulation was set to be 90% of threshold. The baseline condition used for normalisation is the sum of brain-only and spinal-only MEPs. ***B***, In one example participant, the catheter electrode was positioned over the C6 dorsal root entry zone (DREZ) of the spinal cord and the Biceps muscle was targeted. Facilitation was strong in the targeted muscle and present in muscles innervated at nearby segments. Shoulder and leg muscles omitted for visualisation purposes. ***C***, In a different participant, the catheter electrode was placed over the C8 DREZ and the APB was targeted. Strong facilitation was present in the target muscle but was also present in ECR and FCR.

We quantified this selectivity, first for individuals and then across participants (Fig. 9). Fig. 9A shows the muscle selectivity for a single participant. The measure of selectivity was calculated as the relative activation of a muscle under each of the conditions. For example a selectivity of 0.75 in the target flexor carpi radialis (FCR) indicates that the FCR contributes 75% of the total MEP AUC across muscles. This measure was repeated with brain only (110% of threshold), spinal only (110% of threshold), and paired stimulation at the optimal latency. In this participant, FCR was the most activated muscle in all conditions, but the selectivity measure of this muscle was larger for the paired condition than for the brain-only and spinal-only conditions.

**Figure 9.**
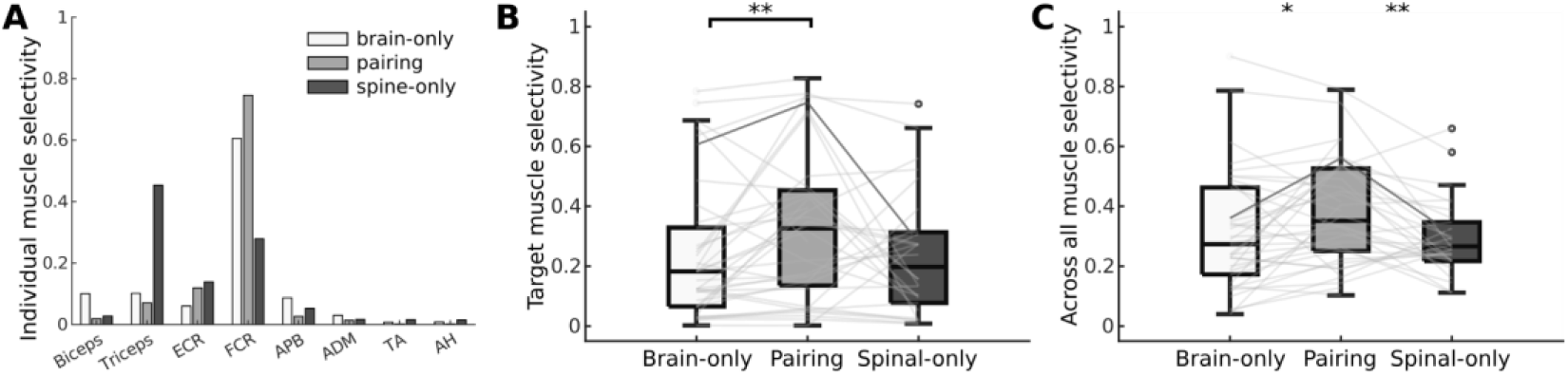
Paired brain and spine stimulation yields more selective activation of individual muscles. *A*, Example of individual muscle selectivity from a single participant showing that for the FCR and APB the selectivity of pairing is larger than for either brain-only or spinal-only stimulation alone. ***B*,** Selectivity as in A can be pooled across participants (n = 38, 23M/15F) by selecting the selectivity corresponding to the target muscle from each participant. Median target muscle selectivity is higher for pairing stimulation than for brain-only stimulation. While it is also higher for pairing stimulation than spinal-only stimulation this difference is not statistically significant. ***C*,** Across muscle selectivity measures the selectivity of muscle activation irrespective of the target muscle and is higher for pairing stimulation than for both brain-only stimulation and spinal-only stimulation (n = 38, 23M/15F). For B and C: Individual lines correspond to individual participants. Dark line corresponds to the participant shown in A. Hinges represent 1st and 3rd quartile, and whiskers span the range of the data not considered outliers (defined as q3 + 1.5 × (q3 − q1) or less than q1 − 1.5 × (q3 − q1)).

The selectivity of muscle activation was compared across all participants. Selectivity in the targeted muscle was larger for paired than for brain-only stimulation (37.5%, *p* = 0.002, n = 38, signed-rank test) but not for spinal-only stimulation (43.5%, *p* = 0.094, n = 30, signed-rank test, Fig. 9B). To ensure that these effects were not due to the choice of muscles, the selectivity of activation was compared across muscles. In order to condense multiple muscle selectivities into a single value, we used the normalised metric of entropy that is small when MEP sizes are similar across muscles and large when a single muscle MEP dominates. Fig. 9C shows that when considering all muscles jointly, paired stimulation produces more selective muscle responses than either brain-only (22.6%, *p* = 0.014, n = 38, signed-rank test) or spinal-only stimulation (31.0%, *p* = 0.003, n = 30, signed-rank test).

We also determined whether selectivity has a dependence on ISI. A change in selectivity over the brain-only and spinal-only conditions in an individual participant is strongest at 7-8 ms in their targeted muscle (Fig. 10A). A maximum average across participant selectivity change in the target muscle of 109.2 ± 49.0% (*p* = 0.013, n = 38, signed-rank test; Fig. 10) is consistent with the optimal ISI of 9 ms. Similarly, the maximum across muscle measure of selectivity is 33.9 ± 15.0% at 9 ms (*p* = 0.022, n = 38, signed-rank test; Fig. 10). The increase in selectivity change towards the optimal ISI is strongly related to a corresponding increase in facilitation (slope = 4.74 ± 0.81, *p* = 2.4×10^-4^, n = 11, Fig. 10). Thus, there is a strong correlation (R^2^ = 0.79) between how much a muscle is facilitated by pairing and how selective is the activation of the target muscle.

**Figure 10.**
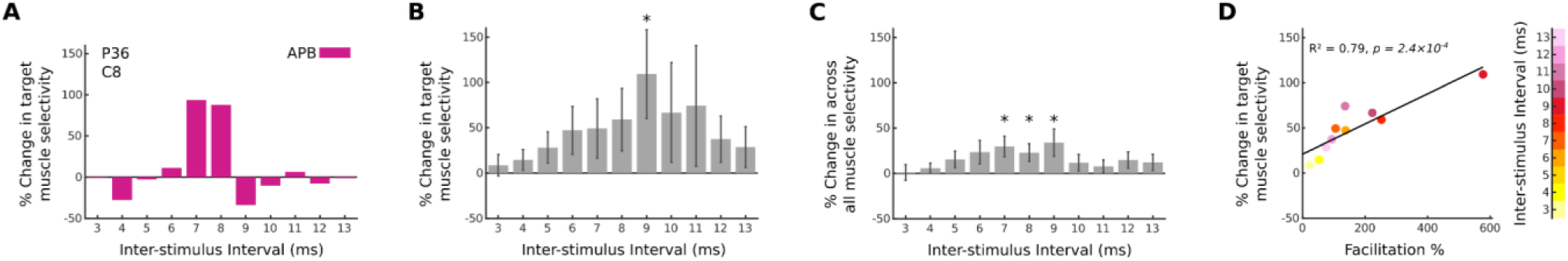
Activation of muscles is most selective near optimum inter-stimulus intervals. *A*, Example of target muscle selectivity (APB) from a single participant. Change in selectivity of pairing from brain- and spinal-only is strongest at an inter-stimulus interval of 7-8 ms. 0% change in selectivity indicates that pairing selectivity is the same as the selectivity computed on the sum of the brain-only (110%) and spinal-only (90%) MEP-size. ***B,*** Selectivity change as in A can be pooled across participants (n = 38, 23M/15F) by choosing the selectivity corresponding to the target muscle from each participant. Average target selectivity is highest at an ISI of 9 ms, corresponding to the timing of optimal pairing. ***C,*** Change in the across muscle selectivity measure also shows the highest selectivity to be at an ISI of 9 ms, albeit at lower magnitude. Across-participant signed-rank test, \**p* < .05, not corrected for multiple comparisons (n = 38, 23M/15F). ***D,*** The selectivity change (as in B) increases (t-test, n = 38, 23M/15F) as the facilitation increases with varying ISI (Fig. 3A).

### 3.7 Degree of impairment does not influence strength of facilitation

In previous work we found that the facilitatory effects of combined brain and spinal stimulation were strong in both uninjured (Mishra et al. 2017) and spinal cord injured (Pal et al. 2022) rats. Consistent with this, there was no relationship between MEP augmentation and radiographic (T2 signal) or clinical evidence of myelopathy (mJOA, reflexes). Specifically, clinical measures of hand use (mJOA; *p* = 0.696; n = 38, Kruskal–Wallis test, Fig. 11A), average strength of the forearm and hand (MRC scale; *p* = 0.375; n = 35, Kruskal–Wallis test, Fig. 11B), a reflex score measured in the biceps muscle (*p* = 0.324; n = 28, Kruskal–Wallis test, Fig. 11C), and a reflex score measured in the triceps muscle (*p* = 0.410, n = 27, Kruskal–Wallis test, Fig. 11D).

**Figure 11.**
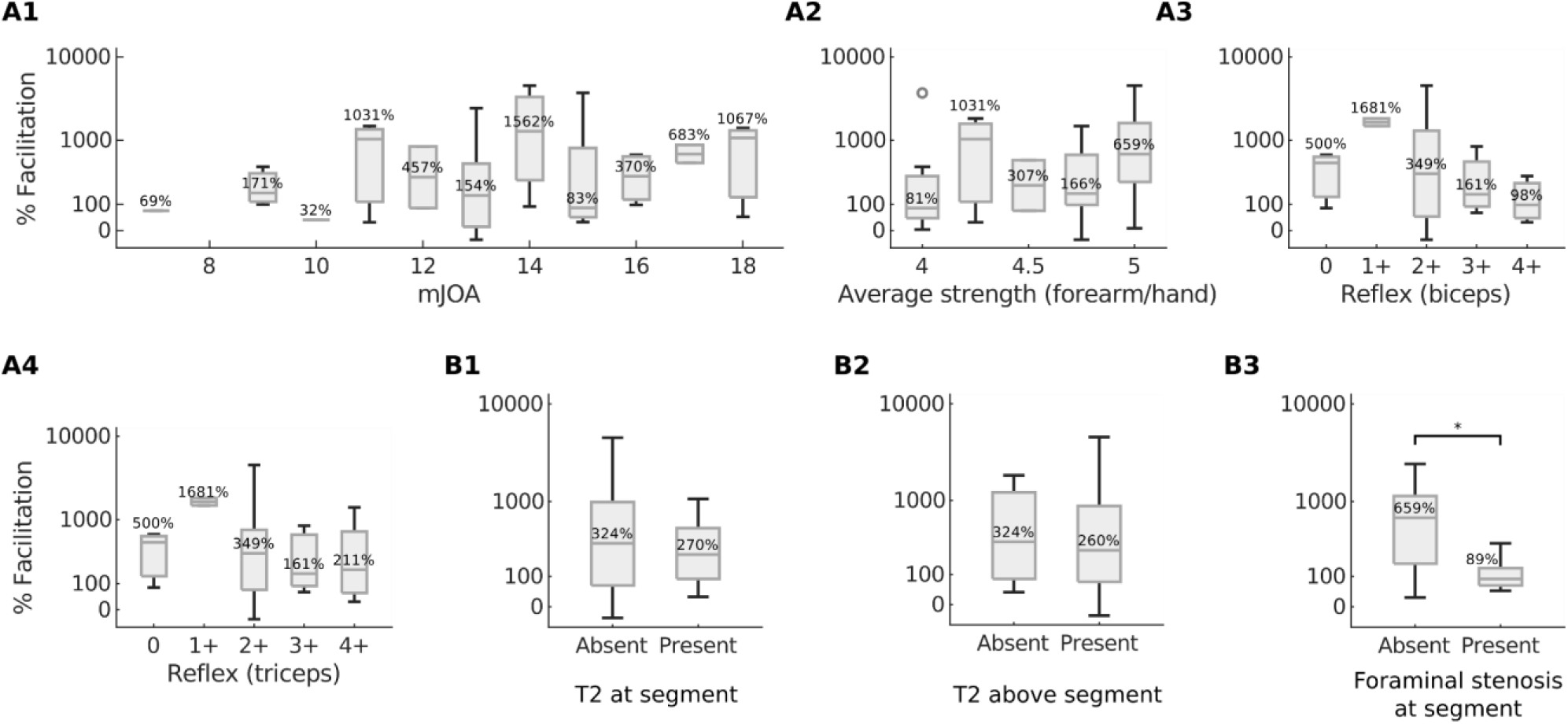
No relation between immediate effect size and degree of impairment. *A1*, mJOA. ***A2***, Average strength in the forearm and hand (MRC scale). ***A3***, Biceps reflex score. ***A4***, Triceps reflex score. ***B1***, Hyperintensity on T2-weighted MRI signal at the stimulated segment. ***B2***, T2 hyperintensity bove the stimulated segment. ***B3***, Severe foraminal stenosis at the stimulated segment. Text represents median % facilitation. Across-participant signed-rank test, \**p* < .05, no correction for multiple comparisons was applied.

Additionally, no relationship was detectable between immediate effect size and the presence of T2-weighted signal change at the segment (*p* = 0.726, n = 37, Wilcoxon rank-sum test; Fig. 11E) or above the segment (*p* = 0.742, n = 37, Wilcoxon rank-sum test; Fig. 11F). However, the presence of severe foraminal stenosis at the stimulated segment was associated with weaker facilitation (*p* = 0.042, n = 22, Wilcoxon rank-sum test; Fig. 11G).

## 4 Discussion

Our understanding of the interactions between descending motor systems and segmental afferents was advanced in four important ways. First, posterior but not anterior spinal cord stimulation augmented cortical MEPs, suggesting that synergistic effects of pairing involve descending motor and spinal sensory interactions. Second, the facilitation induced by spinal stimulation was maximal at the time that motor cortex stimuli arrived in the spinal cord, suggesting this location as the site of interaction. Third, pairing at the optimal inter-stimulus interval made muscle responses more selective, in addition to making them stronger. Fourth, the degree of facilitation was not detectably influenced by radiographic (T2 signal) or clinical evidence of myelopathy (mJOA, reflexes).

We found a synergistic facilitation of MEPs when brain stimulation was combined with stimulation over the posterior– but not anterior–cervical spinal cord. This finding suggests that the interaction critical for the convergence mechanism that we observed takes place at the sensory-motor interface rather than solely within the motor system. This aligns with the understanding that anterior stimulation directly engages the motor unit (Guiho, Baker, and Jackson 2021). In contrast, the observed synergistic facilitation (greater than 5 fold) observed while pairing posterior spinal cord stimulation is consistent with facilitation in rats and monkeys (2-3 fold) that has been demonstrated to be mediated by afferents (Guiho,

Baker, and Jackson 2021; Pal et al. 2022), the primary neural target of SCS (Capogrosso et al. 2013; Greiner et al. 2021; Minassian et al. 2004). Prior research in rats has shown that repeated pairing of the posterior cervical spinal cord and motor cortex at the ISI that produces the largest facilitation (optimal ISI) also leads to lasting plasticity effects (Pal et al. 2022). Nonetheless, the lack of facilitatory effects we observed from combined brain and anterior spinal cord stimulation does not necessarily preclude the induction of plasticity with repeated stimulation that directly engages the motor unit (Bunday and Perez 2012).

The timing of spinal stimulation relative to brain stimulation emerges as a critical factor in realising these synergistic effects. In all tested conditions, the latency of strongest facilitation was observed at 3 ms after the last stimulation pulse; when taken together with the estimate of corticospinal conduction time, this time interval suggests that the synergistic interaction is occurring in the cervical spinal cord. In a single participant, a central conduction time of 2.8 ms was estimated with an epidural spinal cord recording in response to brain stimulation (Di Lazzaro and Rothwell 2014). In this participant, the onset of facilitation was observed to be 1 - 1.3 ms. If this facilitation onset and the central conduction time were equal, it would imply that the spinal stimulation produced instant convergence, however the difference between these two values of 1.5 ms (Mills and Murray 1986; Taylor and Martin 2009) suggests the presence of a single synaptic delay.

The paired conditions are more selective than the brain- or spinal-only conditions. This increase in selectivity appears to be directly coupled to the inter-stimulus interval dependent increase in facilitation. It is currently unclear whether neuromodulation strategies are more effective when they are highly targeted to individual muscles (high selectivity) or when they are weakly targeted (low selectivity) producing broad activation of multiple muscles. Selective activation might benefit patient recovery without causing excessive muscle activation or off-target effects. When optimised, pairing brain and spinal stimulation may allow for more targeted plasticity in muscles that are clinically important to strengthen.

Synergistic effects of pairing brain and spinal stimulation were present regardless of the severity of myelopathy as measured by clinical signs or spinal cord imaging. This finding is consistent with our previous observation that pairing effects that were equally strong in uninjured rats and rats with spinal cord injury (Mishra et al. 2017; Pal et al. 2022), which spares <1% of the corticospinal tract (Yang et al. 2019). The presence of severe foraminal stenosis was associated with weaker pairing effects. Since the facilitation is mediated by the presence of afferent fibres, this observation is consistent with H-reflex suppression observed in radiculopathy (Mazzocchio et al. 2001), potentially caused by axonal loss of afferents. We demonstrated that spinal stimulation can be a potent modulator of weak corticospinal activation, resonating with human studies where volitional control was regained despite severe damage to the neural pathway (Angeli et al. 2018; Carhart et al. 2004; Gill et al. 2018; Inanici et al. 2018; 2021; Powell et al. 2023; Rowald et al. 2022; Wagner et al. 2018). With regards to delivering paired brain and spinal stimulation as therapy, as has been applied in rats (Pal et al. 2022), it is also noteworthy that synergistic effects can be accessed at low intensities that may be tolerated by patients. In short, the independence of the synergistic effect from the degree of myelopathy suggests that repeated pairing could be viable as therapy for spinal cord injury.

Despite the large effect size on average, some participants did not display any synergistic effect. While the presence of severe foraminal stenosis at the stimulated segment may be associated with weaker facilitation, there may be other factors that contribute to this variability. As we have shown by comparing anterior and posterior stimulation, the position of the electrode is critical. For example, while large diameter afferents are the lowest threshold fibres in response to posterior cord stimulation (Greiner et al. 2021), it has been demonstrated that the corticospinal tract can be activated when it is specifically targeted (Vedran Deletis et al. 2018; V. Deletis and Bueno De Camargo 2001). Thus, deviations in the location of the electrodes may contribute to the activation of alternative tracts or fibres. Despite confirmation of electrode position via clinically indicated imaging in a subset of cases, the precise location of the electrode could not be directly visually inspected because it was placed under the lamina. Further study with intraoperative imaging and more targeted electrode configurations will be needed to understand the relationship between position and efficacy of stimulation.

The limitations of this study are largely related to the physiology being performed during a clinically indicated surgery. Experiments were performed under general anaesthesia, which can alter responses. However, while it is conceivable that general anaesthesia specifically affects the synergistic effect of paired stimulation, in rat studies synergistic effects were observed both in anaesthetised (Mishra et al. 2017) and awake animals (Pal et al. 2022). The surgery also limited the segments of the spinal cord that could be stimulated. The catheter electrode was placed at one segment below the most caudal laminectomy, and only one segment was tested in each surgery. Because of this, the most common segments tested were C7-T1, and the upper arm was targeted for pairing in less than 15% of experiments. Despite increased selectivity with pairing, we observed some synergistic effects at muscles innervated at distant segments. Due to intraoperative constraints, we were unable to record full recruitment curves of the synergistic, brain-only and spinal-only conditions, or determine their saturation points.

This study paves the way to test the repeated application of combining brain and spinal stimulation in humans with the aim to invoke spinal cord associative plasticity mechanisms that have been previously observed in rats (Pal et al. 2022). The goal will be to convert the immediate changes in MEP size observed in this study into adaptive changes in the sensorimotor system that persist, taking advantage of the spinal cord’s capacity for plasticity (Wolpaw 2010). We will also explore whether paired stimulation requires epidural electrodes or whether non-invasive methods could be used (Gad et al. 2018; Hofstoetter and Minassian 2022). This research deepens our understanding of spinal cord stimulation and may inform neurorehabilitation intervention based on associative stimulation of brain and spinal cord in humans.

## 5.3 Acknowledgements

We thank neurologists A. Mendiratta, P. Kent, H. Choi, M. Bell (The Och Spine Hospital At New York Presbyterian Hospital), S. C. Karceski (Weill Cornell Medicine) and intraoperative monitoring technologists N. Patel, Z. Moheet (Weill Cornell Medicine), Joe Elliott, Brian Demboski, Kelley Wichman, Susannah Storms, Meghan Mullaney, Evance Desriviere (The Och Spine Hospital At New York Presbyterian Hospital) for monitoring patient safety during the experiments, as well as help with running the experiments. We also thank M. Vulapalli, C. Mykolajtchuk, J. Berger, (Weill Cornell Medicine), J. Reyes and P. Martinez (The Och Spine Hospital At New York Presbyterian Hospital) for help with patient recruitment and administrative matters.

## 5.4 Author contributions

Conceptualization: JRM, NYH, MSV, JBC; Data Curation: JRM, PG; Formal Analysis: JRM, JBC; Funding Acquisition: JRM, NYH, MSV, JBC; Investigation: JRM, EFJ, JLG, PG, JML, ZMS, RAL, AKC, KDR, MSV, CM, JBC; Methodology: JRM, ET, OM, ES, NYH, MSV, CM, JBC; Project Administration: JRM, MSV, JBC; Resources: ET, OM, ES, ZMS, RAL, AKC, KDR, NYH, MSV, CM, JBC; Software: JRM; Supervision: ET, ES, MSV, JBC; Validation: JRM, LM, NYH, MSV, JBC; Visualisation: JRM, LM, NYH, JBC; Writing – Original Draft: JRM, JBC; Writing – Review & Editing: JRM, EFJ, JLG, PG, LM, ET, OM, ES, JML, ZMS, RAL, AKC, KDR, NYH, MSV, CM, JBC.

## 5.5 Data availability statement

The data that support the findings of this study are available from the corresponding authors, upon reasonable request.

## 6 Supporting Information

*Supporting Table 1* - Tested experimental conditions.

*Supporting Table 2* - Detailed clinical characteristics of participants.

*Supporting Figure 1* - Convergence of posterior subthreshold spinal stimulation and suprathreshold brain stimulation in individual participants.

*Supporting Figure 2* - Convergence of anterior subthreshold spinal stimulation and suprathreshold brain stimulation in individual participants.

## Abbreviations

ADM: abductor digiti minimi
AH: Abductor Hallucis
APB: abductor pollicis brevis
AUC: Area Under the Curve
DREZ: dorsal root entry zone
ECR: extensor carpi radialis
EDB: Extensor Digitorum Brevis
SCS: spinal cord stimulation
FCR: flexor carpi radialis
mJOA: Modified Japanese Orthopaedic Association
MEP: Motor Evoked Potential
SEM: Standard Error of the Mean
TA: Tibialis Anterior
tES: transcranial electrical stimulation.

## 8 Supporting information

### 8.1 Tested experimental conditions

**Supporting Information Table 1.** Tested experimental conditions.

|  | Posterior, Triple-pulse<br>tES (110%) – SCS (90%) | Posterior, Triple-pulse<br>tES (110%) – SCS (110%) | Posterior, Single-pulse<br>tES (<100%) – SCS (110%) | Anterior, Triple-pulse<br>tES (110%) – SCS (90%) | Anterior, Triple-pulse<br>tES (110%) – SCS (90%) |
| --- | --- | --- | --- | --- | --- |
| P01 | C8R-APB | C8R-APB |  |  |  |
| P02 | C8L-APB |  |  |  |  |
| P03 | C8L-APB | C8L-APB |  |  |  |
| P04 | C7R-Triceps |  |  |  |  |
| P05 | C8L-ADM |  |  |  |  |
| P06 | C8R-APB |  |  |  |  |
| P07 |  |  |  | C7R-Triceps | C7R-Triceps |
| P08 | C8L-APB |  |  |  |  |
| P09 | T1R-ADM |  |  |  |  |
| P10 | C8R-APB |  |  |  |  |
| P11 |  |  |  | C8R-APB |  |
| P12 |  |  |  | C6L-Biceps | C6L-Biceps |
| P13 |  |  |  | C7R-APB | C7R-APB |
| P14 |  |  |  | T1R-APB |  |
| P15 | C8R-Biceps |  |  |  |  |
| P16 | C6R-APB |  |  |  |  |
| P17 | C7L-Biceps |  |  |  |  |
| P18 |  |  |  | C8R-APB |  |
| P19 | C8L-ADM |  |  | C7L-ADM |  |
| P20 | C7R-APB |  |  |  |  |
| P21 | C5L-Triceps |  |  |  |  |
| P22 |  |  |  | C8L-APB | C8L-APB |
| P23 | C7R-Triceps |  |  |  |  |
| P24 | C6R-Biceps |  |  |  |  |
| P25 | C7R-APB |  |  |  |  |
| P26 | C8L-APB |  |  |  |  |
| P27 | C8L-APB |  |  |  |  |
| P28 |  |  |  | C8R-APB |  |
| P29 | C8L-APB |  |  |  |  |
| P30 | C7L-Triceps |  |  |  |  |
| P31 | C7L-APB |  |  |  |  |
| P32 | C7R-APB | C7R-FCR |  |  |  |
| P33 | C7L-FCR | C7L-FCR |  |  |  |
| P34 | C7L-ADM |  |  |  |  |
| P35 | C7R-ADM |  |  |  |  |
| P36 | C8L-APB |  |  |  |  |
| P37 | C8L-APB |  |  |  |  |
| P38 | C8L-APB | C8L-APB |  |  |  |
| P39 | C8L-APB |  |  |  |  |
| P40 |  | C8L-APB | C8L-APB |  |  |
| P41 | C8R-APB | C8R-APB | C8R-APB |  |  |
| P42 | C8R-APB | C8L-APB |  |  |  |
| P43 | C8R-APB |  |  |  |  |
| P44 | C8L-APB | C8L-APB | C8L-APB |  |  |
| P45 |  | C7L-APB | C7L-APB |  |  |
| P46 | T1R-APB | T1R-FCR | T1R-FCR |  |  |
| P47 |  |  | C8L-FCR |  |  |
| P48 |  | C7R-APB | C7R-FCR |  |  |
| P49 | C8L-APB | C8L-APB | C8L-APB |  |  |
| P50 | T1R-FCR | T1R-FCR | T1R-FCR |  |  |
| P51 |  |  |  | C8L-APB | C8L-APB |
| P52 |  |  |  |  | C6L-ECR |
| P53 |  |  |  | C8L-APB | C8L-APB |
| P54 |  | C8L-APB | C8L-APB |  |  |
| P55 | C8L-FCR |  |  |  |  |
| P56 |  |  | C7R-FCR |  |  |
| P57 |  | C8R-ECR |  |  |  |
| P58 |  |  | C7L-FCR |  |  |
| P59 |  |  |  | C8R-APB | C8R-APB |
| Total: | 38 | 10 | 11 | 12 | 8 |
| For each participant, the site of epidural spinal cord stimulation (SCS), side of the body (L/R), and name of the target muscle, is listed for each paired condition tested. Percentages refer to threshold relative to the target muscle MEP. ADM, abductor digiti minimi; APB, abductor pollicis brevis; ECR, extensor carpi radialis; FCR, flexor carpi radialis. |  |  |  |  |  |

### 8.2 Detailed clinical characteristics of participants

**Supporting Information Table 2.**
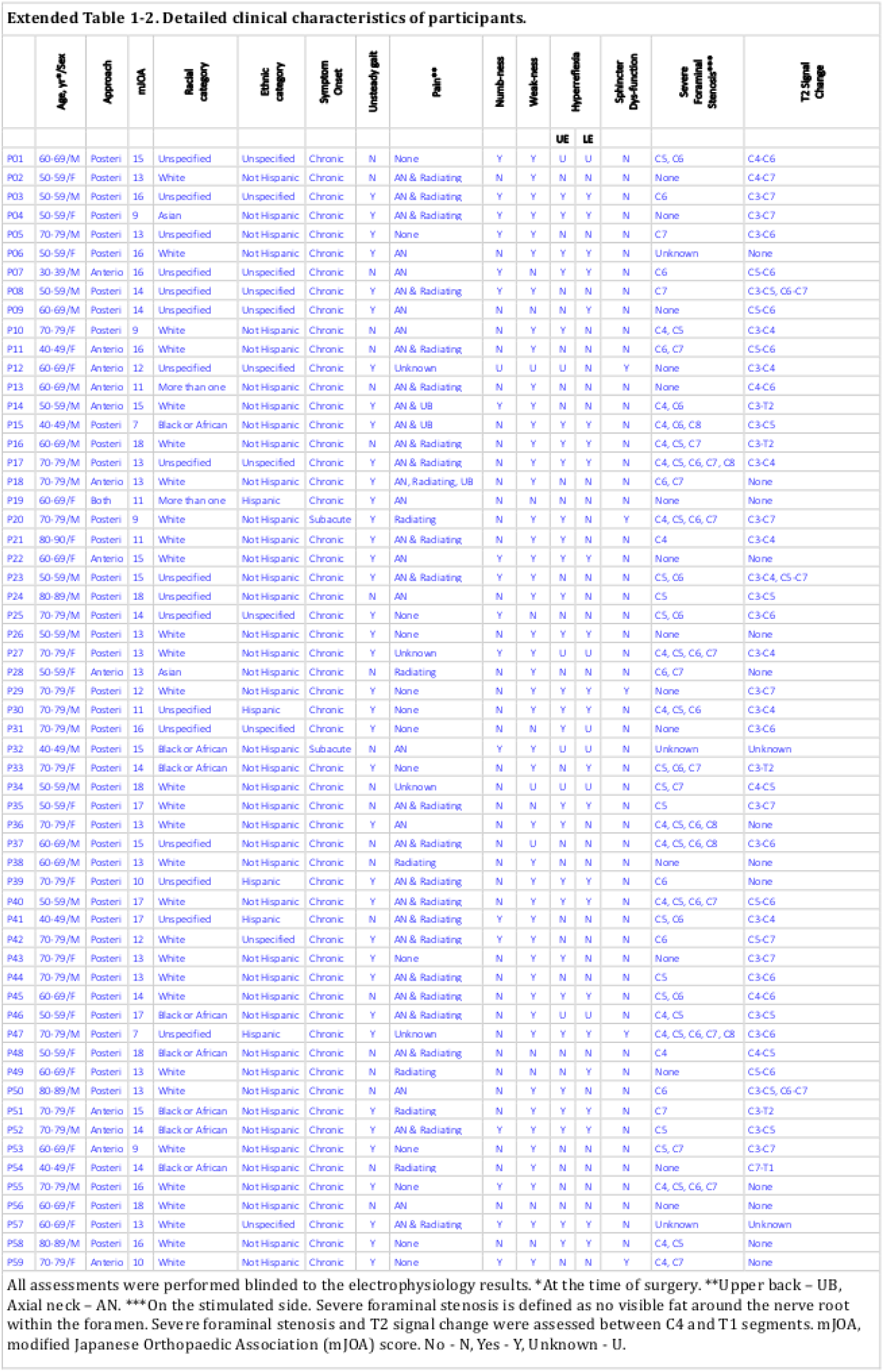
Detailed clinical characteristics of participants.

### 8.3 Convergence of posterior subthreshold spinal stimulation and suprathreshold brain stimulation in individual participants

**Supporting Information Figure 1.**
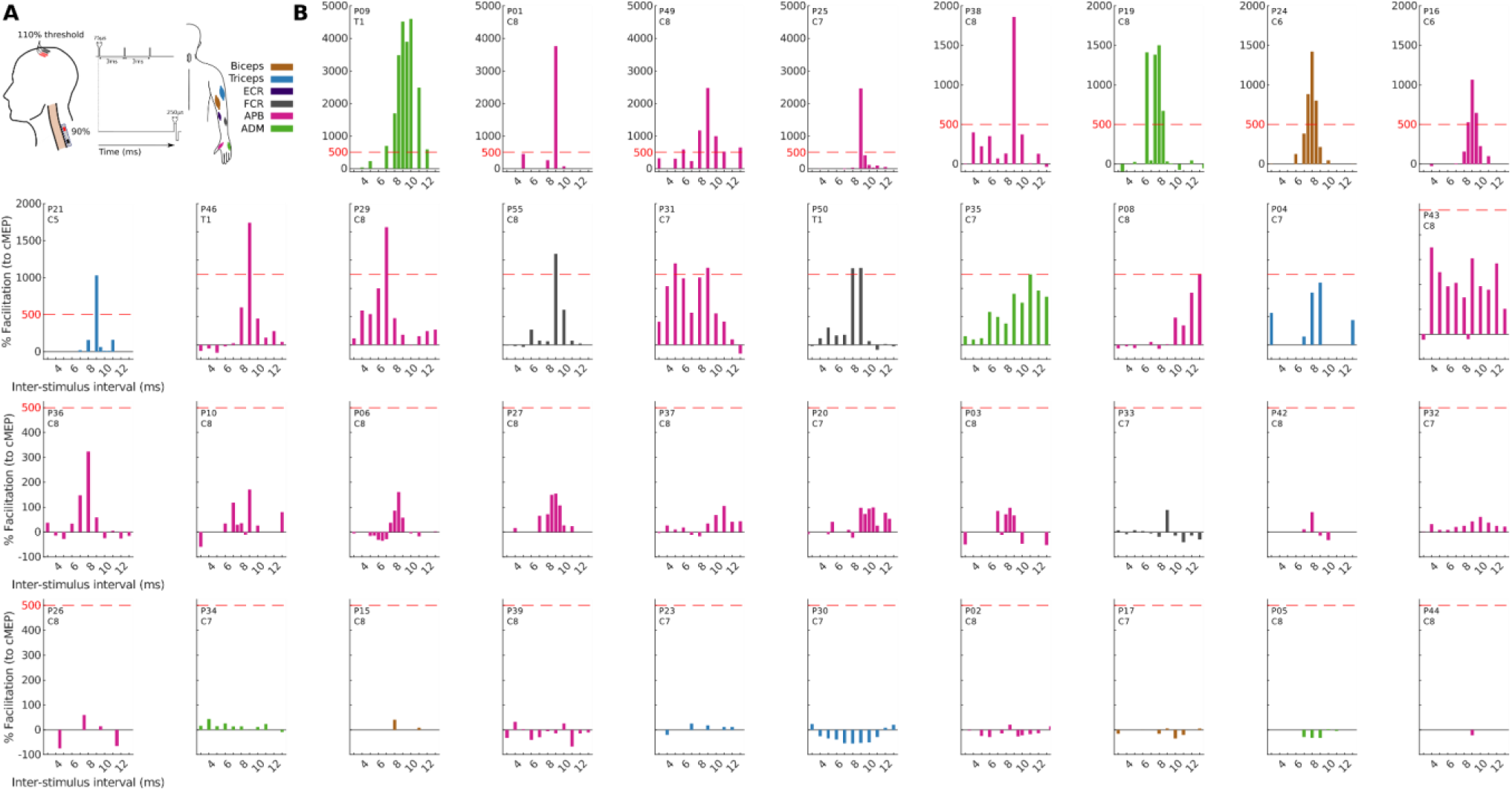
Posterior subthreshold spinal stimulation converges with suprathreshold brain stimulation in individual participants. *A*, Schematic diagram representing the triple-pulse sequence delivered to the brain and the single pulse delivered to the spinal cord. The catheter electrode was positioned at different segments in different participants targeting the dorsal root entry zone (DREZ) of the spinal cord. ***B***, When the catheter is placed on the dorsal aspect of the dura targeting the DREZ (n = 38, 23M/15F), a strong facilitation is visible in the majority of participants in the range 8-10ms. In a subset of participants, facilitation appears absent. The facilitation is calculated relative to the brain-only motor evoked potential (MEP) size, with a 0% facilitation indicating that the MEP observed in the paired condition is the same size as the MEP in the brain-only condition. Bar chart ordering is sorted by maximum facilitation and color indicates the plotted targeted muscle (legend shown in A). P36 is duplicated from Fig. 2. ADM, abductor digiti minimi; APB, abductor pollicis brevis; ECR, extensor carpi radialis; FCR, flexor carpi radialis.

### 8.4 Convergence of anterior subthreshold spinal stimulation and suprathreshold brain stimulation in individual participants

**Supporting Information Figure 2.**
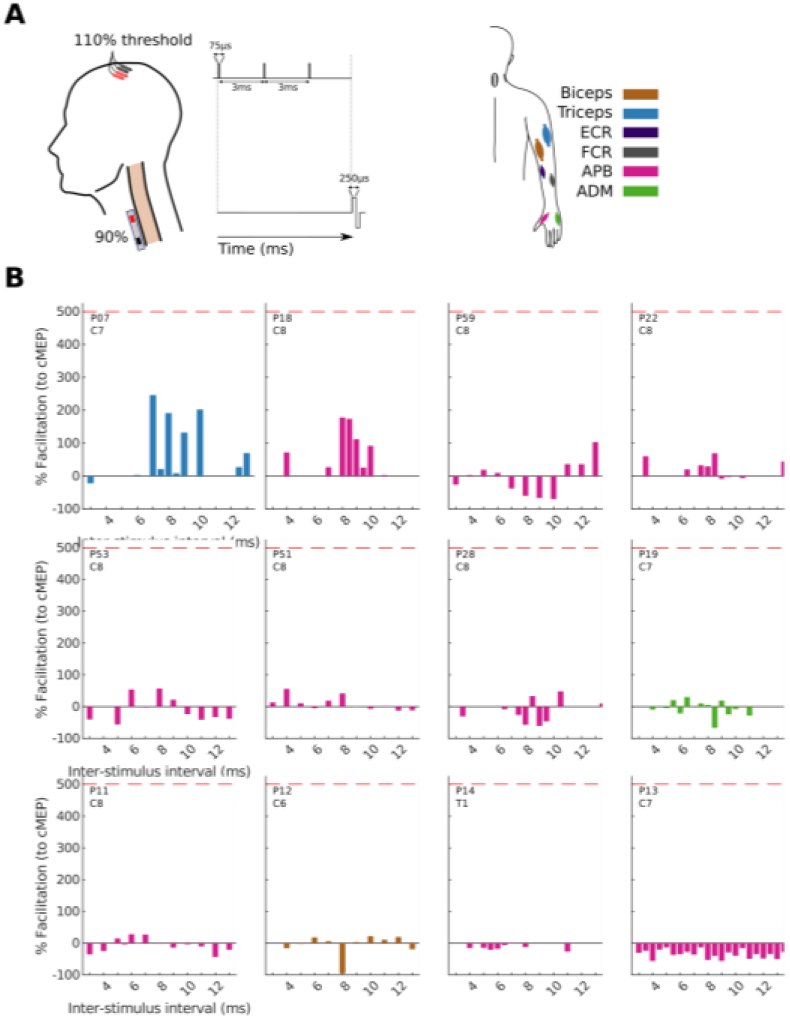
Anterior subthreshold spinal stimulation converges with suprathreshold brain stimulation in individual participants. *A*, Schematic diagram representing the triple-pulse sequence delivered to the brain and the single pulse delivered to the spinal cord. The catheter electrode was positioned at different segments in different participants targeting the ventral root exit zone of the spinal cord. ***B***, When the catheter is placed on the anterior aspect of the dura (n = 12, 4M/8F), facilitation is not present in the majority of participants. The facilitation is calculated relative to the brain-only motor evoked potential (MEP) size, with a 0% facilitation indicating that the MEP observed in the paired condition is the same size as the MEP in the brain-only condition. Bar chart ordering is sorted by maximum facilitation and color indicates the plotted targeted muscle (legend shown in A). ADM, abductor digiti minimi; APB, abductor pollicis brevis; ECR, extensor carpi radialis; FCR, flexor carpi radialis.

## References

Al’joboori, Yazi, Ricci Hannah, Francesca Lenham, Pia Borgas, Charlotte J. P. Kremers, Karen L. Bunday, John Rothwell, and Lynsey D. Duffell. 2021. “The Immediate and Short-Term Effects of Transcutaneous Spinal Cord Stimulation and Peripheral Nerve Stimulation on Corticospinal Excitability.” Frontiers in Neuroscience 15. https://www.frontiersin.org/articles/10.3389/fnins.2021.749042.

Angeli, Claudia A., Maxwell Boakye, Rebekah A. Morton, Justin Vogt, Kristin Benton, Yangshen Chen, Christie K. Ferreira, and Susan J. Harkema. 2018. “Recovery of Over-Ground Walking after Chronic Motor Complete Spinal Cord Injury.” New England Journal of Medicine 379 (13): 1244–50. 10.1056/NEJMoa1803588.

Asan, Ahmet S., James R. McIntosh, and Jason B. Carmel. 2022. “Targeting Sensory and Motor Integration for Recovery of Movement After CNS Injury.” Frontiers in Neuroscience 15. https://www.frontiersin.org/articles/10.3389/fnins.2021.791824.

Benzel, Edward C., John Lancon, Lee Kesterson, and Theresa Hadden. 1991. “Cervical Laminectomy and Dentate Ligament Section for Cervical Spondylotic Myelopathy.” Clinical Spine Surgery 4 (3): 286.

Brendler, Samuel J. 1968. “The Human Cervical Myotomes: Functional Anatomy Studied at Operation.” Journal of Neurosurgery 28 (2): 105–11. 10.3171/jns.1968.28.2.0105.

Bunday, Karen L., and Monica A. Perez. 2012. “Motor Recovery after Spinal Cord Injury Enhanced by Strengthening Corticospinal Synaptic Transmission.” Current Biology 22 (24): 2355–61. 10.1016/j.cub.2012.10.046.

Bunday, Karen L., M.A. Urbin, and Monica A. Perez. 2018. “Potentiating Paired Corticospinal-Motoneuronal Plasticity after Spinal Cord Injury.” Brain Stimulation 11 (5): 1083–92. 10.1016/j.brs.2018.05.006.

Capogrosso, Marco, Nikolaus Wenger, Stanisa Raspopovic, Pavel Musienko, Janine Beauparlant, Lorenzo Bassi Luciani, Grégoire Courtine, and Silvestro Micera. 2013. “A Computational Model for Epidural Electrical Stimulation of Spinal Sensorimotor Circuits.” Journal of Neuroscience 33 (49): 19326–40. 10.1523/JNEUROSCI.1688-13.2013.

Carhart, M.R., Jiping He, R. Herman, S. D’Luzansky, and W.T. Willis. 2004. “Epidural Spinal-Cord Stimulation Facilitates Recovery of Functional Walking Following Incomplete Spinal-Cord Injury.” IEEE Transactions on Neural Systems and Rehabilitation Engineering 12 (1): 32–42. 10.1109/TNSRE.2003.822763.

Compston, Alastair. 2010. “Aids to the Investigation of Peripheral Nerve Injuries. Medical Research Council: Nerve Injuries Research Committee. His Majesty’s Stationery Office: 1942; Pp. 48 (Iii) and 74 Figures and 7 Diagrams; with Aids to the Examination of the Peripheral Nervous System. By Michael O’Brien for the Guarantors of Brain. Saunders Elsevier: 2010; Pp. [8] 64 and 94 Figures.” Brain 133 (10): 2838–44. 10.1093/brain/awq270.

Cowan, J M, B L Day, C Marsden, and J C Rothwell. 1986. “The Effect of Percutaneous Motor Cortex Stimulation on H Reflexes in Muscles of the Arm and Leg in Intact Man.” The Journal of Physiology 377 (1): 333–47. 10.1113/jphysiol1986.sp016190.

Deletis, V., and A. Bueno De Camargo. 2001. “Interventional Neurophysiological Mapping during Spinal Cord Procedures.” Stereotactic and Functional Neurosurgery 77 (1–4): 25–28. 10.1159/000064585.

Deletis, Vedran, Francesco Sala, and Sedat Ulkatan. 2012. Transcranial Electrical Stimulation and Intraoperative Neurophysiology of the Corticospinal Tract. Edited by Charles M. Epstein, Eric M. Wassermann, and Ulf Ziemann. Vol. 1. Oxford University Press. 10.1093/oxfordhb/9780198568926.013.0008.

Deletis, Vedran, Kathleen Seidel, Francesco Sala, Andreas Raabe, Darko Chudy, Juergen Beck, and Karl F Kothbauer. 2018. “Intraoperative Identification of the Corticospinal Tract and Dorsal Column of the Spinal Cord by Electrical Stimulation.” *Journal of Neurology*, Neurosurgery & Psychiatry 89 (7): 754–61. 10.1136/jnnp-2017-317172.

Deuschl, G., R. Michels, A. Berardelli, E. Schenck, M. Inghilleri, and C. H. Lücking. 1991. “Effects of Electric and Magnetic Transcranial Stimulation on Long Latency Reflexes.” Experimental Brain Research 83 (2): 403–10. 10.1007/BF00231165.

Di Lazzaro, Vincenzo, and John C. Rothwell. 2014. “Corticospinal Activity Evoked and Modulated by Non-Invasive Stimulation of the Intact Human Motor Cortex.” The Journal of Physiology 592 (19): 4115–28. 10.1113/jphysiol.2014.274316.

Gad, Parag, Sujin Lee, Nicholas Terrafranca, Hui Zhong, Amanda Turner, Yury Gerasimenko, and V. Reggie Edgerton. 2018. “Non-Invasive Activation of Cervical Spinal Networks after Severe Paralysis.” Journal of Neurotrauma 35 (18): 2145–58. 10.1089/neu.2017.5461.

Gill, Megan L., Peter J. Grahn, Jonathan S. Calvert, Margaux B. Linde, Igor A. Lavrov, Jeffrey A. Strommen, Lisa A. Beck, et al. 2018. “Neuromodulation of Lumbosacral Spinal Networks Enables Independent Stepping after Complete Paraplegia.” Nature Medicine 24 (11): 1677–82. 10.1038/s41591-018-0175-7.

Greiner, Nathan, Beatrice Barra, Giuseppe Schiavone, Henri Lorach, Nicholas James, Sara Conti, Melanie Kaeser, et al. 2021. “Recruitment of Upper-Limb Motoneurons with Epidural Electrical Stimulation of the Cervical Spinal Cord.” Nature Communications 12 (1): 435. 10.1038/s41467-020-20703-1.

Guiho, Thomas, Stuart N. Baker, and Andrew Jackson. 2021. “Epidural and Transcutaneous Spinal Cord Stimulation Facilitates Descending Inputs to Upper-Limb Motoneurons in Monkeys.” Journal of Neural Engineering 18 (4). 10.1088/1741-2552/abe358.

Guzmán-López, Jessica, João Costa, Aikaterini Selvi, Gonzalo Barraza, Jordi Casanova-Molla, and Josep Valls-Solé. 2012. “The Effects of Transcranial Magnetic Stimulation on Vibratory-Induced Presynaptic Inhibition of the Soleus H Reflex.” Experimental Brain Research 220 (3): 223–30. 10.1007/s00221-012-3131-7.

Harris, Paul A., Robert Taylor, Brenda L. Minor, Veida Elliott, Michelle Fernandez, Lindsay O’Neal, Laura McLeod, et al. 2019. “The REDCap Consortium: Building an International Community of Software Platform Partners.” Journal of Biomedical Informatics 95 (July): 103208. 10.1016/j.jbi.2019.103208.

Harris, Paul A., Robert Taylor, Robert Thielke, Jonathon Payne, Nathaniel Gonzalez, and Jose G. Conde. 2009. “Research Electronic Data Capture (REDCap)—A Metadata-Driven Methodology and Workflow Process for Providing Translational Research Informatics Support.” Journal of Biomedical Informatics 42 (2): 377–81. 10.1016/j.jbi.2008.08.010.

Hofstoetter, Ursula S., and Karen Minassian. 2022. “Transcutaneous Spinal Cord Stimulation: Advances in an Emerging Non-Invasive Strategy for Neuromodulation.” Journal of Clinical Medicine 11 (13): 3836. 10.3390/jcm11133836.

Inanici, Fatma, Lorie N. Brighton, Soshi Samejima, Christoph P. Hofstetter, and Chet T. Moritz. 2021. “Transcutaneous Spinal Cord Stimulation Restores Hand and Arm Function After Spinal Cord Injury.” IEEE Transactions on Neural Systems and Rehabilitation Engineering 29: 310–19. 10.1109/TNSRE.2021.3049133.

Inanici, Fatma, Soshi Samejima, Parag Gad, V. Reggie Edgerton, Christoph P. Hofstetter, and Chet T. Moritz. 2018. “Transcutaneous Electrical Spinal Stimulation Promotes Long-Term Recovery of Upper Extremity Function in Chronic Tetraplegia.” IEEE Transactions on Neural Systems and Rehabilitation Engineering 26 (6): 1272–78. 10.1109/TNSRE.2018.2834339.

Jackson, Andrew, Jaideep Mavoori, and Eberhard E. Fetz. 2006. “Long-Term Motor Cortex Plasticity Induced by an Electronic Neural Implant.” Nature 444 (7115): 56–60. 10.1038/nature05226.

Knikou, Maria. 2014. “Transpinal and Transcortical Stimulation Alter Corticospinal Excitability and Increase Spinal Output.” PLoS ONE 9 (7): e102313. 10.1371/journal.pone.0102313.

Lehky, Sidney R., Terrence J. Sejnowski, and Robert Desimone. 2005. “Selectivity and Sparseness in the Responses of Striate Complex Cells.” Vision Research 45 (1): 57–73. 10.1016/j.visres.2004.07.021.

Liang, Lucy, Arianna Damiani, Matteo Del Brocco, Evan R. Rogers, Maria K. Jantz, Lee E. Fisher, Robert A. Gaunt, Marco Capogrosso, Scott F. Lempka, and Elvira Pirondini. 2023. “A Systematic Review of Computational Models for the Design of Spinal Cord Stimulation Therapies: From Neural Circuits to Patient-Specific Simulations.” The Journal of Physiology 601 (15): 3103–21. 10.1113/JP282884.

Mazzocchio, Riccardo, Giovanni Battista Scarfò, Aldo Mariottini, Vitaliano Francesco Muzii, and Lucio Palma. 2001. “Recruitment Curve of the Soleus H-Reflex in Chronic Back Pain and Lumbosacral Radiculopathy.” BMC Musculoskeletal Disorders 2 (October): 4. 10.1186/1471-2474-2-4.

McIntosh, James R., Evan F. Joiner, Jacob L. Goldberg, Lynda M. Murray, Bushra Yasin, Anil Mendiratta, Steven C. Karceski, et al. 2023. “Intraoperative Electrical Stimulation of the Human Dorsal Spinal Cord Reveals a Map of Arm and Hand Muscle Responses.” Journal of Neurophysiology 129 (1): 66–82. 10.1152/jn.00235.2022.

Mills, K. R., and N. M. F. Murray. 1986. “Electrical Stimulation over the Human Vertebral Column: Which Neural Elements Are Excited?” Electroencephalography and Clinical Neurophysiology 63 (6): 582–89. 10.1016/0013-4694(86)90145-8.

Minassian, K., B. Jilge, F. Rattay, M. M. Pinter, H. Binder, F. Gerstenbrand, and M. R. Dimitrijevic. 2004. “Stepping-like Movements in Humans with Complete Spinal Cord Injury Induced by Epidural Stimulation of the Lumbar Cord: Electromyographic Study of Compound Muscle Action Potentials.” Spinal Cord 42 (7): 401–16. 10.1038/sj.sc.3101615.

Mishra, Asht M., Ajay Pal, Disha Gupta, and Jason B. Carmel. 2017. “Paired Motor Cortex and Cervical Epidural Electrical Stimulation Timed to Converge in the Spinal Cord Promotes Lasting Increases in Motor Responses.” The Journal of Physiology 595 (22): 6953–68. 10.1113/JP274663.

Nishimura, Yukio, Steve I. Perlmutter, Ryan W. Eaton, and Eberhard E. Fetz. 2013. “Spike-Timing-Dependent Plasticity in Primate Corticospinal Connections Induced during Free Behavior.” Neuron 80 (5): 1301–9. 10.1016/j.neuron.2013.08.028.

Pal, Ajay, HongGeun Park, Aditya Ramamurthy, Ahmet S Asan, Thelma Bethea, Meenu Johnkutty, and Jason B Carmel. 2022. “Spinal Cord Associative Plasticity Improves Forelimb Sensorimotor Function after Cervical Injury.” *Brain*, September, awac235. 10.1093/brain/awac235.

Poon, David E., Francois D. Roy, Monica A. Gorassini, and Richard B. Stein. 2008. “Interaction of Paired Cortical and Peripheral Nerve Stimulation on Human Motor Neurons.” Experimental Brain Research 188 (1): 13–21. 10.1007/s00221-008-1334-8.

Powell, Marc P., Nikhil Verma, Erynn Sorensen, Erick Carranza, Amy Boos, Daryl P. Fields, Souvik Roy, et al. 2023. “Epidural Stimulation of the Cervical Spinal Cord for Post-Stroke Upper-Limb Paresis.” Nature Medicine 29 (3): 689–99. 10.1038/s41591-022-02202-6.

Rowald, Andreas, Salif Komi, Robin Demesmaeker, Edeny Baaklini, Sergio Daniel Hernandez-Charpak, Edoardo Paoles, Hazael Montanaro, et al. 2022. “Activity-Dependent Spinal Cord Neuromodulation Rapidly Restores Trunk and Leg Motor Functions after Complete Paralysis.” *Nature Medicine*, February, 1–12. 10.1038/s41591-021-01663-5.

Roy, François D., Dillen Bosgra, and Richard B. Stein. 2014. “Interaction of Transcutaneous Spinal Stimulation and Transcranial Magnetic Stimulation in Human Leg Muscles.” Experimental Brain Research 232 (6): 1717–28. 10.1007/s00221-014-3864-6.

Seeman, Stephanie C., Brian J. Mogen, Eberhard E. Fetz, and Steve I. Perlmutter. 2017. “Paired Stimulation for Spike-Timing-Dependent Plasticity in Primate Sensorimotor Cortex.” The Journal of Neuroscience 37 (7): 1935–49. 10.1523/JNEUROSCI.2046-16.2017.

Stefan, Katja, Erwin Kunesch, Leonardo G. Cohen, Reiner Benecke, and Joseph Classen. 2000. “Induction of Plasticity in the Human Motor Cortex by Paired Associative Stimulation.” Brain 123 (3): 572–84. 10.1093/brain/123.3.572.

Szelényi, Andrea, Karl F. Kothbauer, and Vedran Deletis. 2007. “Transcranial Electric Stimulation for Intraoperative Motor Evoked Potential Monitoring: Stimulation Parameters and Electrode Montages.” Clinical Neurophysiology 118 (7): 1586–95. 10.1016/j.clinph.2007.04.008.

Taylor, Janet L., and Peter G. Martin. 2009. “Voluntary Motor Output Is Altered by Spike-Timing-Dependent Changes in the Human Corticospinal Pathway.” Journal of Neuroscience 29 (37): 11708–16. 10.1523/JNEUROSCI.2217-09.2009.

Wagner, Fabien B., Jean-Baptiste Mignardot, Camille G. Le Goff-Mignardot, Robin Demesmaeker, Salif Komi, Marco Capogrosso, Andreas Rowald, et al. 2018. “Targeted Neurotechnology Restores Walking in Humans with Spinal Cord Injury.” Nature 563 (7729): 65–71. 10.1038/s41586-018-0649-2.

Wolpaw, Jonathan R. 2010. “What Can the Spinal Cord Teach Us about Learning and Memory?” The Neuroscientist 16 (5): 532–49. 10.1177/1073858410368314.

Wolpert, Daniel M., Jörn Diedrichsen, and J. Randall Flanagan. 2011. “Principles of Sensorimotor Learning.” Nature Reviews. Neuroscience 12 (12): 739–51. 10.1038/nrn3112.

Yang, Qi, Aditya Ramamurthy, Sophia Lall, Joshua Santos, Shivakeshavan Ratnadurai-Giridharan, Madeleine Lopane, Neela Zareen, et al. 2019. “Independent Replication of Motor Cortex and Cervical Spinal Cord Electrical Stimulation to Promote Forelimb Motor Function after Spinal Cord Injury in Rats.” Experimental Neurology 320 (October): 112962. 10.1016/j.expneurol.2019.112962.

